# High-Resolution Digital Phenotypes from Consumer Wearables Enhance Prediction of Cardiometabolic Risk Markers

**DOI:** 10.1101/2021.10.29.21265547

**Authors:** Weizhuang Zhou, Yu En Chan, Chuan Sheng Foo, Jingxian Zhang, Jing Xian Teo, Sonia Davila, Weiting Huang, Jonathan Yap, Stuart A. Cook, Patrick Tan, Calvin Woon-Loong Chin, Khung Keong Yeo, Weng Khong Lim, Pavitra Krishnaswamy

**Affiliations:** Institute for Infocomm Research, Agency for Science Technology and Research (A*STAR), Singapore; SingHealth Duke-NUS Institute of Precision Medicine, Singapore; SingHealth Duke-NUS Genomic Medicine Centre, Singapore; Cardiovascular and Metabolic Disorders Program, Duke-NUS Medical School, Singapore, Singapore; Department of Cardiology, National Heart Centre Singapore, Singapore; Duke-NUS Medical School, Singapore; Cancer and Stem Biology Program, Duke-NUS Medical School, Singapore; Cancer Science Institute of Singapore, National University of Singapore, Singapore; Genome Institute of Singapore, Agency for Science Technology and Research (A*STAR), Singapore

**Keywords:** Wearable device, heart rate, cardiometabolic disease, risk prediction, digital phenotypes, polygenic risk scores, time series analysis, machine learning, free-living

## Abstract

**Background:** Consumer-grade wearable devices enable detailed recordings of heart rate and step counts in free-living conditions. Recent studies have shown that summary statistics from these wearable recordings have potential uses for longitudinal monitoring of health and disease states. However, the relationship between higher resolution physiological dynamics from wearables and known markers of health and disease remains largely uncharacterized.

**Objective:** We aimed to (i) derive high resolution digital phenotypes from observational wearable recordings, (ii) characterize their ability to predict modifiable markers of cardiometabolic disease, and (iii) study their connections with genetic predispositions for cardiometabolic disease and with lifestyle factors.

**Methods:** We introduce a principled framework to extract interpretable high resolution phenotypes from wearable data recorded in free-living conditions. The proposed framework standardizes handling of data irregularities, encodes contextual information about underlying physiological state at any given time, and generates a set of 66 minimally redundant features across active, sedentary and sleep states. We applied our approach on a multimodal dataset, from the SingHEART study (NCT02791152), that comprises of heart rate and step count time series from wearables, clinical screening profiles, whole genome sequences and lifestyle survey responses from 692 healthy volunteers. We employed machine learning to model non-linear relationships between the high resolution phenotypes and clinical risk markers for blood pressure, lipid and weight abnormalities. For each risk type, we performed model comparisons based on Brier Skill Scores (BSS) to assess predictive value of the high resolution features over and beyond typical baselines. We then examined associations between the wearable-derived features, polygenic risk for cardiometabolic disease, and lifestyle habits and health perceptions.

**Results:** Compared to typical summary statistic measures like resting heart rate, we find that the high-resolution features collectively have greater predictive value for modifiable clinical markers associated with cardiometabolic disease risk (average improvement in Brier Skill Score=52.3%, *P*<.001). Further, we show that heart rate dynamics from different activity states contain distinct information about type of cardiometabolic risk, with dynamics in sedentary states being most predictive of lipid abnormalities and patterns in active states being most predictive of blood pressure abnormalities (*P*<.001). Finally, our results reveal that subtle heart rate dynamics in wearable recordings serve as physiological correlates of genetic predisposition for cardiometabolic disease, lifestyle habits and health perceptions.

**Conclusions:** High resolution digital phenotypes recorded by consumer wearables in free-living states have the potential to enhance prediction of cardiometabolic disease risk, and could enable more proactive and personalized health management.

**Trial Registration:** ClinicalTrials.gov NCT02791152; https://clinicaltrials.gov/ct2/show/NCT02791152

## Introduction

The adoption of consumer-grade wearable activity trackers into routine use has been increasing rapidly in recent years, with approximately one in five U.S. adults reported to regularly use wrist-worn smartwatches and fitness trackers in 2019 [1]. This phenomenon has generated an unprecedented scale of consumer health data, and led to many studies on the wider health uses of such data. These studies are increasingly generating evidence to reveal relations between recordings from wearable activity trackers and the risk for conditions ranging from mental health and infectious disease [2,3] to cardiovascular and metabolic (“cardiometabolic”) diseases [4–7]. Amongst these, due to apparent links between activity levels and cardiometabolic health, the evidence for broader health uses of wearables is most established in the cardiometabolic domain [4,8–11].

Previous studies in the cardiometabolic domain have focused on the utility of wearable-derived summary statistics, and fall into one of two categories. First, electrocardiogram signals from wearables have been studied in relation to the development of cardiometabolic conditions such as atrial fibrillation [12–14], hyperkalaemia [15,16] and heart failure [17–19]. As many of these conditions are amenable to early intervention via dietary changes or increased physical activity, there is also interest in using wearables to promote self-awareness and regulation [20] and to enhance screening [11]. Second, wearable-derived measures, such as circadian measures, sleep patterns/quality [11,21], step counts [4], wearable-derived resting heart rate [4,8,10,21,22] and heart rate variability (HRV) [23–27] have been found to correlate with outcomes in cardiometabolic disease. As such, there is increasing recognition in the clinical community to incorporate wearable-derived measures into practical cardiometabolic disease management [6,28].

At the same time, consumer wearable technology is developing rapidly and enabling better, richer, and more granular measurements. In particular, advanced sensors in present day consumer wearable and mobile health devices enable recording and extraction of step counts, activity patterns, heart rate, and sleep states at fine temporal resolutions [6,29,30]. Therefore, the logical evolution of research in this area would be to extract higher-resolution features, beyond traditional summary statistics and standard wearable-derived measurements, from these time series recordings, and to assess their utility in relation to cardiometabolic health states. A few recent studies have employed black-box deep neural networks to relate high-resolution heart rate and step count time series recorded using wearables to the risk of developing atrial fibrillation, sleep apnoea and hypertension [31,32]. However, as their primary goal focused on risk target classification, the nature of the intermediate predictive time series features and their connection with other known biological and lifestyle-related markers of cardiometabolic disease remains unresolved.

In this paper, we introduce a framework to derive interpretable high resolution features from heart rate and step count time series recorded by consumer wearables. We applied our approach on multidimensional data from normal volunteers in the SingHEART study [33] to examine associations of high resolution wearable-based phenotypes with diverse indicators of cardiometabolic health and disease. We found that high resolution digital phenotypes from wearables had higher predictive value for modifiable risk markers of cardiometabolic disease than established summary statistic measures like resting heart rate. In order to better understand the wearable-derived features, we investigated how they relate to genetic predispositions for cardiometabolic disease and to lifestyle habits. We discovered that subtle high resolution patterns in wearable recordings may reflect subclinical physiological changes associated with both genetic risk markers and lifestyle factors. Our findings have implications for use of digital phenotypes from consumer wearables as objective and quantitative indicators of cardiometabolic health and disease.

## Methods

### Data

We sourced data from the SingHEART study (NCT02791152) as of October 8, 2019. This study [33,34] was established at the National Heart Center Singapore (NHCS), a tertiary specialty hospital in Singapore, and approved by the SingHealth Centralized Institutional Review Board (CIRB Ref: 2015/2601 and 2018/3081). Enrolment targeted healthy volunteers who provided written informed consent to use data (including electronic health records) for research. Subjects were required to fulfil the following inclusion criteria: (i) 21-69 y/o, (ii) no personal medical history of prior cardiovascular disease (myocardial infarction (MI), coronary artery disease (CAD), peripheral arterial disease (PAD), stroke), cancer, autoimmune/genetic disease, endocrine disease, diabetes mellitus, psychiatric illness, asthma, chronic lung disease or chronic infective disease, and (iii) no family medical history of cardiomyopathies.

At the point of enrolment, each subject was profiled using a range of health assessment modalities. The resulting dataset includes (a) heart rate and step count time series recordings over 3-5 days from consumer wearable devices (Fitbit^®^ Charge HR), together with the associated sleep logs generated by Fitbit^®^, (b) self-reported answers to a lifestyle and quality-of-life questionnaire [4], (c) genotypic data from whole genome sequencing (WGS) using the Illumina HiSeq X platform, and (d) laboratory measurements for nine clinically-relevant markers (systolic and diastolic blood pressure; blood levels of triglycerides, total cholesterol, high density lipoprotein (HDL) and low density lipoprotein (LDL); fasting blood glucose level; waist circumference and body mass index (BMI)). As of October 8, 2019, the full study cohort contained 1,101 subjects, amongst whom 692 subjects had complete wearable recordings. A full description of the data is provided in Supplementary Information (SI) Table 1.

**Table 1:** Description of the Different Model Types

| Model Name | Features Included | Number of Features |
| --- | --- | --- |
| Baseline [4] | Age + Gender | 2 |
| RHR | Baseline features + RestingHR | 3 |
| SummaryStats | Baseline features + Wearable Summary Stats | 12 |
| HighRes.ActiveSeg | Baseline features + Catch22 (active) | 24 |
| HighRes.SedenSeg | Baseline features + Catch22 (sedentary) | 24 |
| HighRes.SleepSeg | Baseline features + Catch22 (sleep) | 24 |

While all health assessments above were performed at the point of enrolment into the study, we also longitudinally tracked each subject for occurrence of any actual clinical events. In particular, we extracted all ICD-10 codes pertaining to any acute care utilization events in the regional health system associated with the NHCS until January 2021 to characterize links between data features, risk markers and actual clinical events.

### Extraction of Features from Wearable Time Series Recordings

We now describe steps to derive resting heart rate, summary statistics on activity and sleep patterns, and high resolution features from the wearable heart rate and step count time series recordings. As all these physiologic features are derived from the same recordings, they are internally consistent and can be meaningfully used for downstream comparative analyses.

#### Computation of Resting Heart Rate

We used the wearable heart rate time series recordings to derive resting heart rate (RestingHR) [4]. Specifically, we defined RestingHR as the average of heart rate values across all time points that had a valid heart rate record and a step count of <=100. We note that there are similarities between wearable-derived resting heart rate and the clinical gold standard, ECG-derived heart rate [4,35].

#### Annotation of Wearable Time Series Recordings

We extracted the wearable time series recordings for each subject, and utilized only days with at least 20 hours of step count and/or heart rate data as per Lim et al [4]. This procedure yielded 642 subjects. Heart rate recordings were available either at regular one-minute intervals, or as irregular bursts of recordings over 5, 10 or 15-second intervals. Step count recordings were sampled at either 15-minute or one-minute intervals. We resampled all heart rate and step count consumer wearable records to one-minute intervals, and then annotated the time series to reflect data availability and physical activity levels (Figure 1A). We assigned a “null” value for heart rate at time points where it was missing. Then, we annotated time points with available data for both heart rate and step count as “sleep”, “active” or “sedentary”. Specifically, we applied the “sleep” annotation to all time points captured by the Fitbit sleep log, the “sedentary” annotation to any time points with zero step count value, and denoted the remaining time points as “active”. On average, the 642 subjects in our study had 3.72 days of valid heart rate data, and the average missing heart rate periods in a day were 94.9 minutes long. The median lengths of the longest uninterrupted time series for the “active”, “sedentary” and “sleep” periods were 31mins, 105mins, and 465mins respectively.

**Figure 1.**
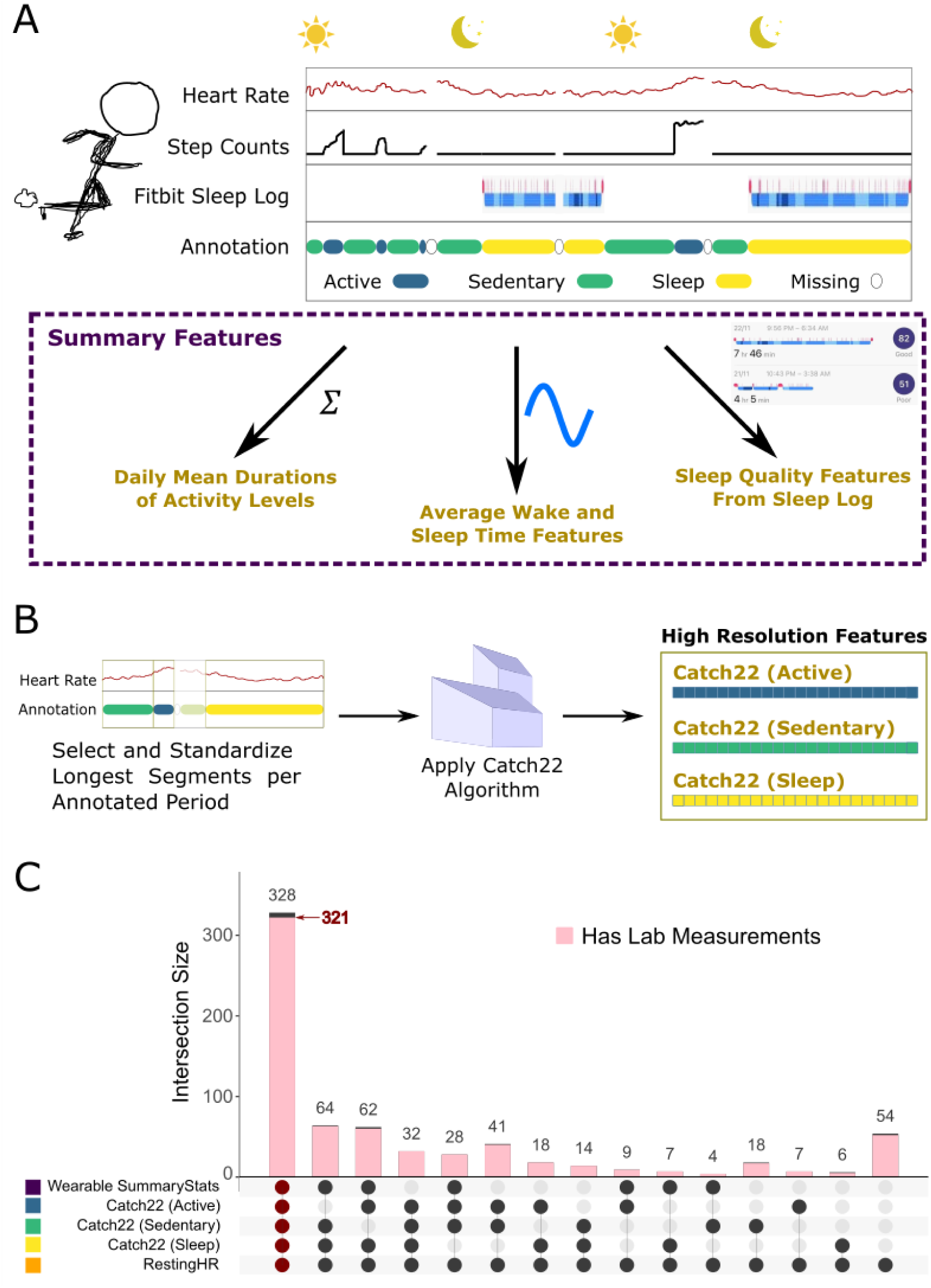
Wearable Data Processing Pipeline. (A) Construction of low-resolution features based on summary statistics. (B) Construction of high-resolution features based on Catch22 algorithm. (C) UpSet plot of the 692 subjects with features from the various categories. Only non-empty set intersections are presented. Intersection size indicates the number of subjects found within the intersections of given sets. Of the largest intersection with 328 subjects, 321 also had laboratory measurement recordings.

For each subject, we processed the heart rate and step count time series recordings from the consumer wearable devices to yield a range of summary and high-resolution features, as detailed below.

#### Derivation of Summary Features from Wearable Time Series Recordings

We employed a three-step procedure to derive a range of wearable summary statistics (Figure 1A). First, we used our physical activity annotations to compute mean daily durations for the different activity levels. Second, we used device logs to obtain statistics relating to sleep-wake patterns. Third, we converted the wake and sleep time into a 24-hr format, and averaged the resulting values over all days where a given subject had wearable data recordings. To account for the cyclical nature of sleep/wake patterns, we transformed the average wake and sleep times using sinusoidal functions. Overall, this process yields 10 summary features for each subject. All the summary statistics included are listed in SI Table 2.

**Table 2:** Lab Measurements and Corresponding Thresholds

|  | Lab Measurement | Threshold to be considered at risk |
| --- | --- | --- |
| I | Systolic Blood Pressure | More than 140 mmHg |
| II | Diastolic Blood Pressure | More than 90 mmHg |
| III | Triglycerides | More than 2.3 mmol/l |
| IV | Total Cholesterol | More than 6.2 mmol/l |
| V | HDL | Less than 1 mmol/l |
| VI | LDL | More than 4.1 mmol/l |
| VII | Fasting Blood Glucose Level | More than 6 mmol/l |
| VIII | Waist Circumference | More than 100 cm (males)/ 90 cm (females) |
| IX | Body Mass Index | More than 27.5 |

#### Derivation of High Resolution Features from Wearable Time Series Recordings

We further developed a data processing pipeline to extract high resolution time series features from the wearable device heart rate recordings (Figure 1B). Reasoning that heart rate and step count patterns under different physiological states or activity levels could provide distinct insights into cardiovascular health, we sought to derive time series features that encode contextual information about the physiological state or activity level. Specifically, we processed heart rate time series recordings for each of the three physical activity levels (sleep, sedentary, active) separately, as follows.

For each subject, we chose the longest uninterrupted time period of the heart rate time series recordings for each physical activity level. Because the data exhibits significant variability in the lengths of these time periods across subjects, we defined pre-specified lengths to extract standardized sleep, sedentary and active segments. Specifically, we extracted the first twenty minutes for active segments, the first one hour for sedentary segments and the first five hours for sleep segments. If the recordings available for a subject did not fulfil the aforementioned length criteria even with the longest segment for a given activity level, we did not consider that particular activity level for high-resolution analyses. This process yields up to three heart rate time series segments for each subject.

We then processed each of these extracted heart rate time series segments to obtain high resolution features. Given a time series segment, it is possible to employ computational packages such as the highly comparative time series analysis [36,37] and TSFRESH [38,39] to generate thousands of high resolution features. However, such approaches can generate many redundant features and the process of selecting a concise but effective representation is often not straightforward. Recent work [40] introduced a minimally redundant and interpretable set of 22 features, termed Catch-22 features, that have high predictive value across 93 diverse time series classification datasets. For any given time-series segment, the Catch22 feature set captures several important dynamical properties, including autocorrelations (both linear and non-linear), value distributions and fluctuation analysis. We applied the Catch22 methodology [40] to obtain 22 high resolution features for each available heart rate time series segment. Collectively, our pipeline results in up to three sets of 22 high resolution features per subject, namely Catch22 (Sleep), Catch22 (Active), and Catch22 (Sedentary). All the Catch22 features included are listed in SI Table 3.

**Table 3:**
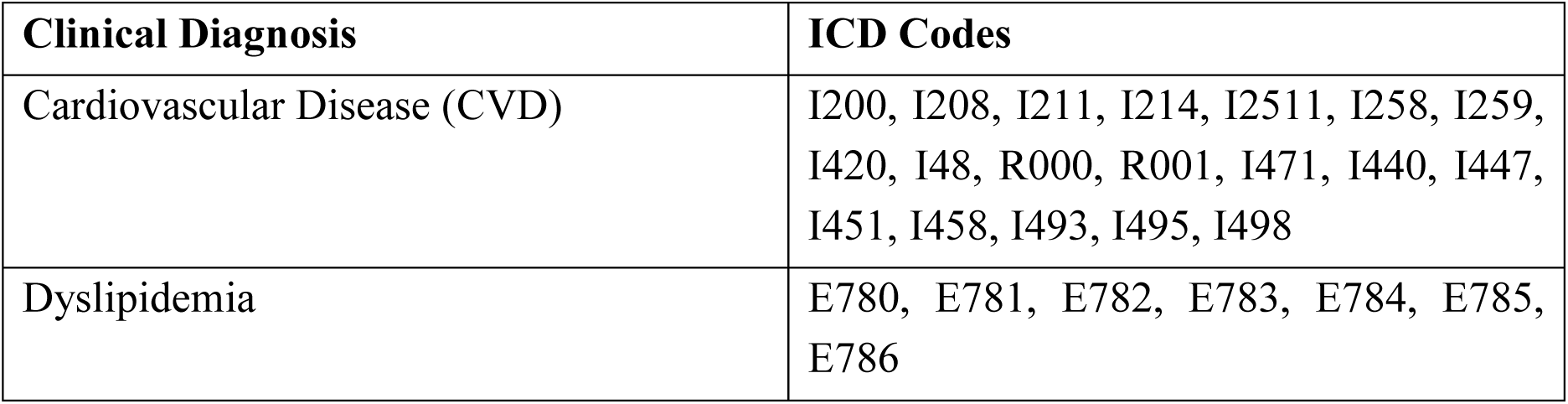
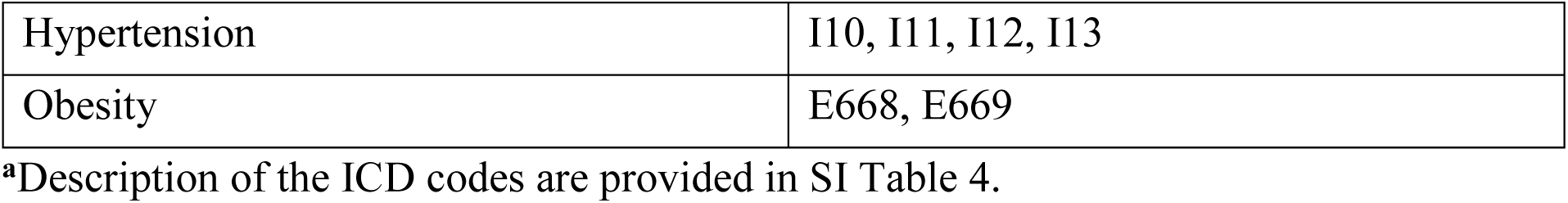
ICD Codes Used for Profiling^**a**^

| Clinical Diagnosis | ICD Codes |
| --- | --- |
| Cardiovascular Disease (CVD) | I200, I208, I211, I214, I2511, I258, I259, I420, I48, R000, R001, I471, I440, I447, I451, I458, I493, I495, I498 |
| Dyslipidemia | E780, E781, E782, E783, E784, E785, E786 |
| Hypertension | I10, I11, I12, I13 |
| Obesity | E668, E669 |

<sup>a</sup>Description of the ICD codes are provided in SI Table 4.

As our study did not prescribe controlled experimental settings for the wearable recordings, the resulting time series segments often exhibit significant noise and irregularities. Hence, we considered the reliability of our featurization approach in these real-world settings. In particular, we assessed stability and sensitivity of the Catch22 features to the length specifications across activity levels (SI-1). The results suggest that the features are relatively robust within the intervals considered, and provide confidence for downstream use of these high resolution features.

#### Overlap amongst Features Derived from Wearable Time Series Recordings

Figure 1C illustrates the overlaps amongst subjects with the different wearable derived features, using UpSet plots [41,42]. For example, 41 individuals had features for active and sedentary segments, but did not have sleep segments or summary statistics (due to lack of sufficiently long continuous sleep recordings). We note that all the different types of wearable features are available for a total of 328 subjects, of which 321 had laboratory measurements as well. We considered this set of 321 subjects for ensuing risk modelling and analysis.

#### Characterization of Predictive Value of Wearable-Derived Features for Clinical Targets

We describe the overall approach to characterize predictive value of the different wearable-derived features with respect to a variety of clinical risk markers. Specifically, we considered model types based on six different feature sets (Table 1). We then defined four target clinical risk markers based on whether the nine laboratory measurements exceeded thresholds in Table 2: (a) abnormal blood pressure (“bp_abnormal”) for either I or II, (b) abnormal lipid levels (“lipids_abnormal”) for at least one of III-VI, (c) obese (“obesity”) for either VIII or IX, and (d) an omnibus category for lipid, blood sugar, obesity and/or sugar abnormalities (“anyRISKoutof9”) for any of I to IX.

All the 321 subjects who had a complete set of wearable-derived features also had complete data for the nine laboratory measurements. We considered this set of 321 subjects as our training set to model clinical risk targets. Of these 321 subjects, 149 were not positive for any of the four risk markers, while 172 were positive for at least one risk marker (see SI-2). We note that a given subject can be positive for more than one of the four labels, but the majority of subjects exhibiting positive risk markers were exclusively labelled by a single risk marker. Specifically, out of the 172 positive subjects, 69% were positive for one clinical risk marker, 23% were positive for two risk markers, and only 8% were positive for three or more risk markers.

We employed machine learning to model the complex non-linear relationships between a given feature set and target pairing using two separate approaches. First, for any given target, we analysed comparative value of the different feature sets (Table 1) by using a model comparison approach. Specifically, we consider the degree to which the wearable-derived features (RestingHR, wearable summary statistics, different high resolution wearable features) augment the predictive value of the baseline demographic feature set, and also compared the performance of the high resolution wearable features against that of the lower resolution features. For appropriate comparison of value-add over the baseline features, all feature sets based on wearable data also include the corresponding baseline feature. Second, for each prediction target, we also compared the importance of the individual feature variables. So as to have a common basis for these variable importance calculations, we developed a unified model with all features included, and used this model to compare variable importance for the different features.

#### Prediction Model and Variable Importance

We trained a series of models to estimate the probability that a subject exhibits clinical risk markers for common cardiometabolic disease abnormalities. Specifically, we used random forest classifiers [43] to model the four targets of interest as they are general purpose non-linear classifiers that perform well in diverse settings. We trained the random forest models in R using the *randomForest* package [44]. To handle the imbalanced nature of the prediction tasks at hand, we set the number of minority class samples chosen for each tree at 80% of the total minority class size. We then downsampled the majority class to match the number of minority class samples used [45]. This was implemented via the *strata* and *sampsize* parameters. For each of the four prediction targets, we constructed 200 such random forests with different starting random seeds, and for each random forest trained, we recorded the out-of-bag (OOB) prediction errors.

To obtain statistically meaningful estimates of variable importance, for a given prediction target, we averaged the mean decrease in accuracy (MDA) for each feature across the 200 random forests. We then ranked the features by their average MDA to obtain the list of top ten ranked features for the target of interest. For analysis of variable importance, we considered the union of the top ten ranking features for the four cardiometabolic disease risk targets.

#### Model Comparison Metric

As our goal is to comparatively characterize predictive value of the different wearable-derived feature sets over and beyond the baseline features, we chose to evaluate relative gains in prediction accuracy. Specifically, as the prediction task is inherently probabilistic, we evaluated the accuracy of probabilistic predictions with the commonly used Brier Skill Score [46,47].

For a given target, we considered the 200 baseline models constructed, computed the Brier Scores for each model (see equation 1), and selected the best performing model with the lowest Brier Score. We denote this selected model as *B*. Then, for each of the other wearable-based model types (Table 1) for the same target, we obtained the corresponding Brier Skill Score as shown in equation 2.

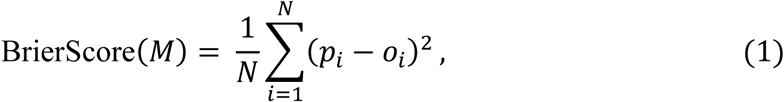

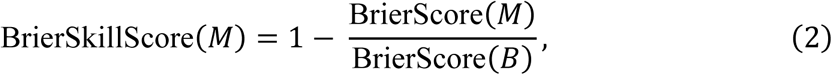

where *M* is the wearable-based model under consideration, *p*_*i*_ is the prediction probability of observing target i using the model under evaluation, *o*_*i*_ is the actual observed target or label (binary: 0/1), and N is the total number of subjects included for modeling. The Brier Skill Score ranges from negative infinity to 1; a positive Brier Skill Score indicates that model *M* performs better than the comparative baseline model *B*, while a negative score indicates model *M* performs worse than the comparative baseline model *B*. If *M* has the exact same performance as *B*, the Brier skill score is zero.

In total, the above process yields 200 Brier Skill Scores for each pairing of prediction target and wearable-derived feature set (model) type. We note that the Skill Scores are based on out-of-bag estimates [43,48], which provides close approximation to an independent test set. We conducted comparisons of Skill Scores between model types via two-sided *t*-tests, with the *P* values adjusted using the Benjamini-Hochberg procedure (BH-adjusted *P* values) [49,50].

#### Illustrative Profiling Based on Clinical Outcomes

Beyond the quantitative characterizations detailed above, we also examined how the high resolution wearable recordings may connect to clinically relevant cardiometabolic disease outcomes. For this, we selected subjects who actualized clinical events with primary ICD10 diagnosis codes for cardiometabolic disease during our longitudinal monitoring period (Table 3). Amongst these subjects, we only considered those not included in the training set for the clinical risk target models. Then, for the selected subjects, we profiled the wearable-derived features alongside other cross-sectional information (demographics, BMI and ECG heart rate).

### Characterization of Interrelations between Wearable-Derived Features, Genetic Risk and Lifestyle Markers

In order to understand the wearable time series features further, we investigated their associations with biological and environmental factors. As probing these associations requires handling diverse multidimensional data types with potentially complex non-linear relationships, we used a machine learning framework (similar to the one described earlier) to construct models of these relationships. We then employed model performance measures to infer the degree of information overlap between the wearable features on the one hand, and (i) genetic risk targets or (ii) lifestyle related targets on the other. For these analyses, we considered the wearable-derived summary statistic features; and the high resolution wearable features with the highest predictive value for the clinical risk markers. We now describe the derivation of the genomic and lifestyle targets, and the setup of the association analyses in each case.

#### Genetic Risk Scoring for Cardiometabolic Diseases

We categorized genetic susceptibility to cardiometabolic diseases using polygenic scores (PGS). As the computation of PGS does not depend on the availability of wearable recording data, we applied the computations to all subjects in our study cohort. The polygenic score catalog [51] is a database of polygenic scores obtained from published scientific studies. As with the NHGRI-EBI GWAS Catalog, the traits corresponding to polygenic scores are grouped by mapped traits [52,53]. To define genetic risk levels for lipid abnormalities, blood pressure abnormalities and obesity, we obtained polygenic scores with less than 20,000 variants from the PGS Catalog based on the mapped trait ontology corresponding to the three targets respectively. For each eligible PGS, we compared the proportion of true cases (based on the laboratory measurements) in the subjects with scores below the 5^th^ percentile and also the subjects with scores above the 95^th^ percentile, and determined the “direction” of the score based on the two proportions. We retained those PGS whose ratio of proportions was >=1.5. This yielded fourteen PGS for lipid abnormalities (PGS000060, PGS000061, PGS000062, PGS000063, PGS000065, PGS000115, PGS000192, PGS000309, PGS000310, PGS000311, PGS000340, PGS000677, PGS000688, PGS000699), two for blood pressure abnormalities (PGS000301, PGS000302) and one for obesity (PGS000298). We detail the scores and mapped trait ontology in SI-3.

For each of the selected PGS, we assigned subjects to high PGS risk and low PGS risk groups if the scores were either larger than the 90^th^ percentile, or smaller than the 10^th^ percentile, depending on the direction of the PGS with respect to the mapped trait propensity. Then, we considered all relevant PGS for a given target (e.g., set of 14 PGS for lipid abnormalities), and labelled subjects with high risk scores for any of the PGS for that trait as having high genetic risk for that trait. For instance, the high genetic risk group for lipid abnormalities would include members with high risk scores for one or more of the fourteen lipid related PGS. The above process provides three binary genetic risk targets – corresponding to high or low genetic risk for lipid abnormalities, blood pressure abnormalities and obesity respectively. In order to evaluate the sensitivity to the chosen percentile cut-offs for genetic risk scores, we repeated analyses for two additional sets of cut-offs: the 80^th^/20^th^ percentile, and 85^th^/15^th^ percentile. We detail the number of subjects for each of the three genetic risk targets under the different cut-offs in SI-4.

We analysed associations between the high-resolution wearable-based physiological features and genetic risk targets. We studied whether the associations with high-resolution features are greater than those with baselines based on gender, resting heart rate, and other summary statistics using model comparison metrics.

#### Lifestyle Habits and Health Perceptions (LH and HP)

We considered a variety of lifestyle habits (LH) and health perceptions (HP) from the LH and HP surveys in our dataset. To choose specific LH and HP targets, we tried to balance data sparsity and diversity as follows. We considered the proportion of subjects who answered the associated questions in the LH and HP surveys, and the diversity in their responses for meaningful analysis. Specifically, we selected only those targets associated with questions that elicited affirmative answers from more than twenty subjects. This resulted in a set of three LH questions, and four HP questions, for a total of seven targets. The targets of the three LH questions focused on (i) consumption of caffeinated drinks, (ii) consumption of alcohol, (iii) adoption of relaxation therapies. The targets of the four HP questions focused on (a) pain/discomfort, (b) anxiety/depression, (c) stress level and (d) health state [4]. We now describe processing choices made to define these targets, and the associated modelling approaches.

For the lifestyle choices, we defined the target Relaxation.Therapies based on the subject’s answer to whether they had engaged in any form of relaxation therapies, and the target alcohol consumption based on whether or not the subject had taken any alcoholic drink within the prior three months to answering the survey. To quantify caffeine intake, we converted the reported weekly consumption of caffeinated beverages into a heuristic score. The beverages taken into consideration were coffee, English tea, Chinese tea and Green tea, and the self-reported levels were “never/rarely”, “less than one cup a week”, “more than or equal to one cup a week, but less than one cup a day”, and “others”. We assigned, for each beverage, scores of 0, 0.5, 2 and 5 respectively for the four levels, and computed the caffeine intake score as the sum of scores for each of the four beverages. We binarized this score and considered a subject as having high caffeine intake if their score was greater than 1.

For the health perception survey, we defined positive and negative classes as follows. For pain/discomfort, we defined the positive class as a choice of “I have moderate pain or discomfort”, and the negative class as a choice of “I have no pain or discomfort”. For anxiety/depression, we defined positive class as a choice of either “I am moderately anxious or depressed” or “I am extremely anxious or depressed”, and the negative class as a choice of “I am not anxious or depressed”. For stress level, as the survey presented subjects with an integer scale of 1 to 10, we divided the answers into three levels: “Low” for scores less than or equal to 3, “Moderate” for scores between 4 and 6 inclusive, and “High” for scores greater than or equal to 7. For Health state, as the survey presented subjects with a continuous scale from 0 to 100, we divided answers into three levels: “Low” for scores less than or equal to 30, “Moderate” for scores greater than 30 but less than or equal to 70, and “High” for scores greater than 70.

We used the same modelling framework as before to assess associations between the wearable-based physiological features and targets relating to lifestyle habits and health perceptions. We studied whether the associations with high-resolution features are greater than those with baselines based on gender and age, resting heart rate, and other summary statistics using model comparison metrics. To choose the subjects for these analyses, we attempted to maximize use of available data and diversity of responses. As responses to the lifestyle questionnaire exhibited higher data sparsity, we chose subjects independently for each modelling target (rather than selecting one fixed set of subjects across all targets). For instance, all models trained to predict alcohol consumption used data from the same set of subjects, regardless of the feature set under consideration. Similarly, all models trained to predict caffeine intake used data from another set of subjects independent of whether the set chosen was identical or intersected with the set used for alcohol consumption. We detail the number of subjects within each target and class in SI-4.

As some of the LH and HP targets contain more than two classes, and sometimes have ordinal values (e.g. low, medium, high), we evaluated the trained models with the more general Ranked Probability Skill Score (RPSS) [54–56] in lieu of the Brier Skill Score. Again, for model *M* and baseline model *B*,

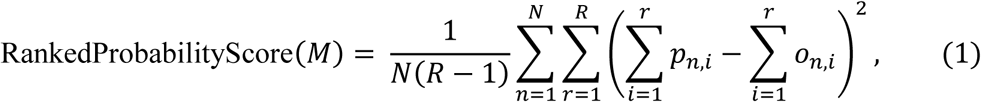

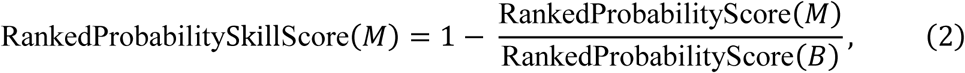

where *p*_*n,i*_ is the predicted cumulative probability of observation *n* for the *i*-th class, *o*_*n,i*_ is the actual cumulative probability for the observation in class *i*-th, *N* is the total number of subjects included in the modelling and *R* is the maximum number of classes in the target parameter.

## Results

### Characteristics of Wearable-Derived High Resolution Heart Rate Features

Unlike summary statistics such as RestingHR which average heart rate measurements across multiple days, our high resolution feature sets constitute a more granular resolution of the heart rate time series dynamics for different physical activity levels. We characterized the different wearable-derived heart rate feature sets by (1) visualizing them as a function of time, and (2) evaluating how they relate to other heart rate features.

First, we examined how the high resolution wearable-derived heart rate features from sleep, active and sedentary segments are distributed across subjects in the study. Figure 2 illustrates the empirical distributions for exemplar features drawn from segments corresponding to each of the three physical activity levels. To examine the variability across subjects, we also visualized representative time series at the 2.5^th^, 25^th^, 50^th^, 75^th^ and 97.5^th^ percentile of the density.

**Figure 2.**
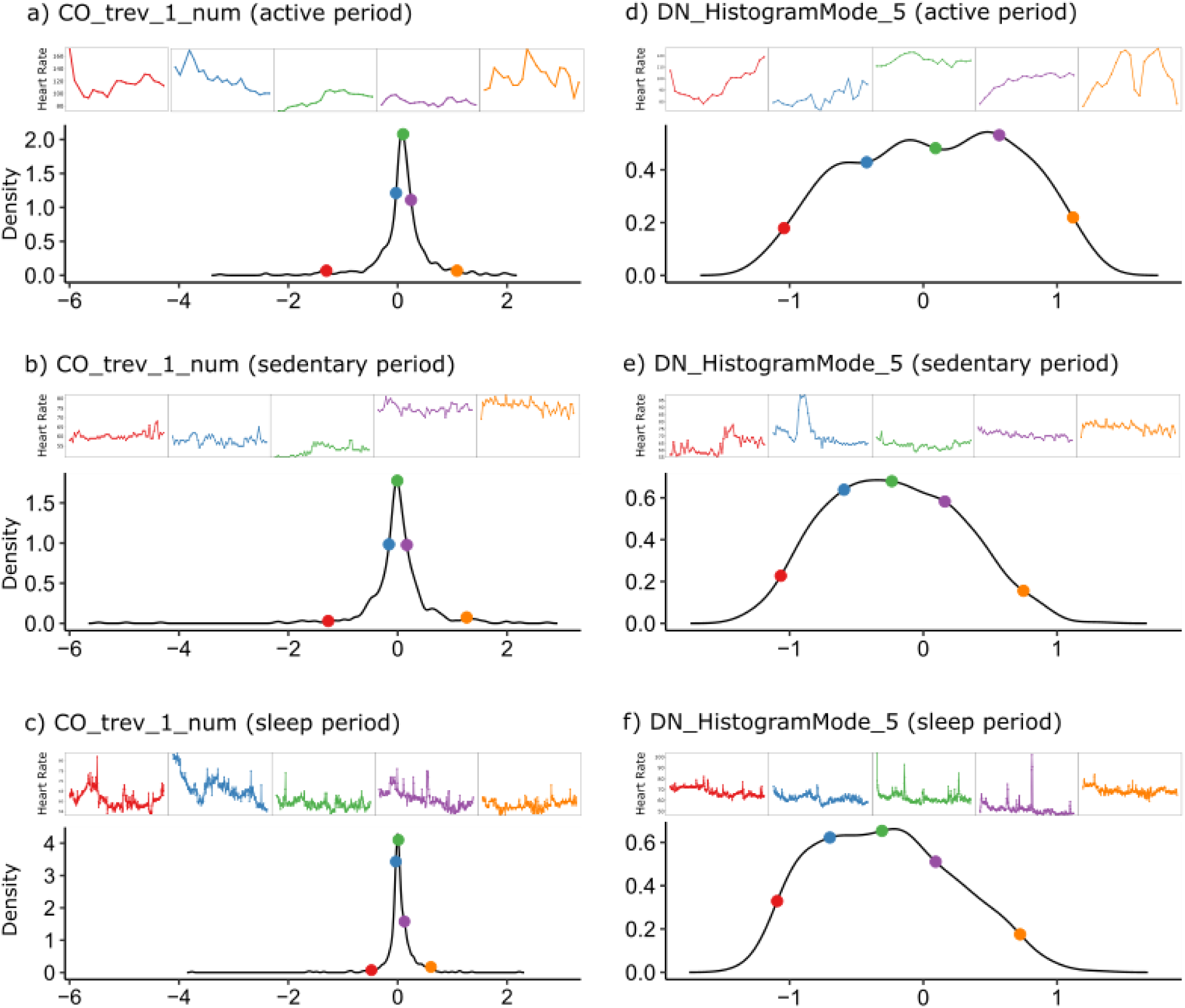
Illustration of Wearable-Derived High-Resolution Heart Rate Features. The distributions of six high-resolution features from the study subjects, based on two Catch22 features obtained from time series recordings in each of the three activity levels. The selected subjects are at the 2.5^th^, 25^th^, 50^th^, 75^th^ and 97.5^th^ percentiles of each distribution, and the time series for the subject is plotted in the corresponding colour. (a-c) CO_trev1_num is the time-reversibility statistic; higher values tend to correspond to more “spiky” and/or irregular time series. (d-f) DN_HistogramMode_5 takes a time series and groups the values across the period into 5 bins, and reports the mode of that graph.

We observe that some features (e.g., the time reversibility statistic ⟨(*x*_*t*+1_ − *x*_*t*_)^3^⟩_*t*_ in Figure 2a-c) relate to the degree of spikiness/regularity in the wearable based heart rate time series, while other features quantify the degree of non-normality of the time series values (e.g. *DN_HistogramMode_5* in Figures 2d-f, which corresponds to the mode of the z-transformed values).

Second, we studied relationships between the different high resolution heart rate features. For any given feature, we considered vectors of feature values for each physical activity level across the population (e.g. CO_trev1_num.active, CO_trev1_num.sedentary), and calculated the angular distance (Figure 3, bottom-right) between feature vectors for each pairing of activity levels. Although the Catch22 algorithm was identically applied to each of the three activity segments, we observed that the angular distances between features generated from the three segments (i.e., for active, sleep, and sedentary states) are generally large (Figure 3A). In some cases, the feature vectors are almost orthogonal to each other (e.g. CO_trev_1_num). We also compared the distributions of Catch 22 feature values across the three different activity levels, and found differences in the distributions (SI-5). These findings suggest that the same feature may contain distinct information about heart rate dynamics in different activity states.

**Figure 3.**
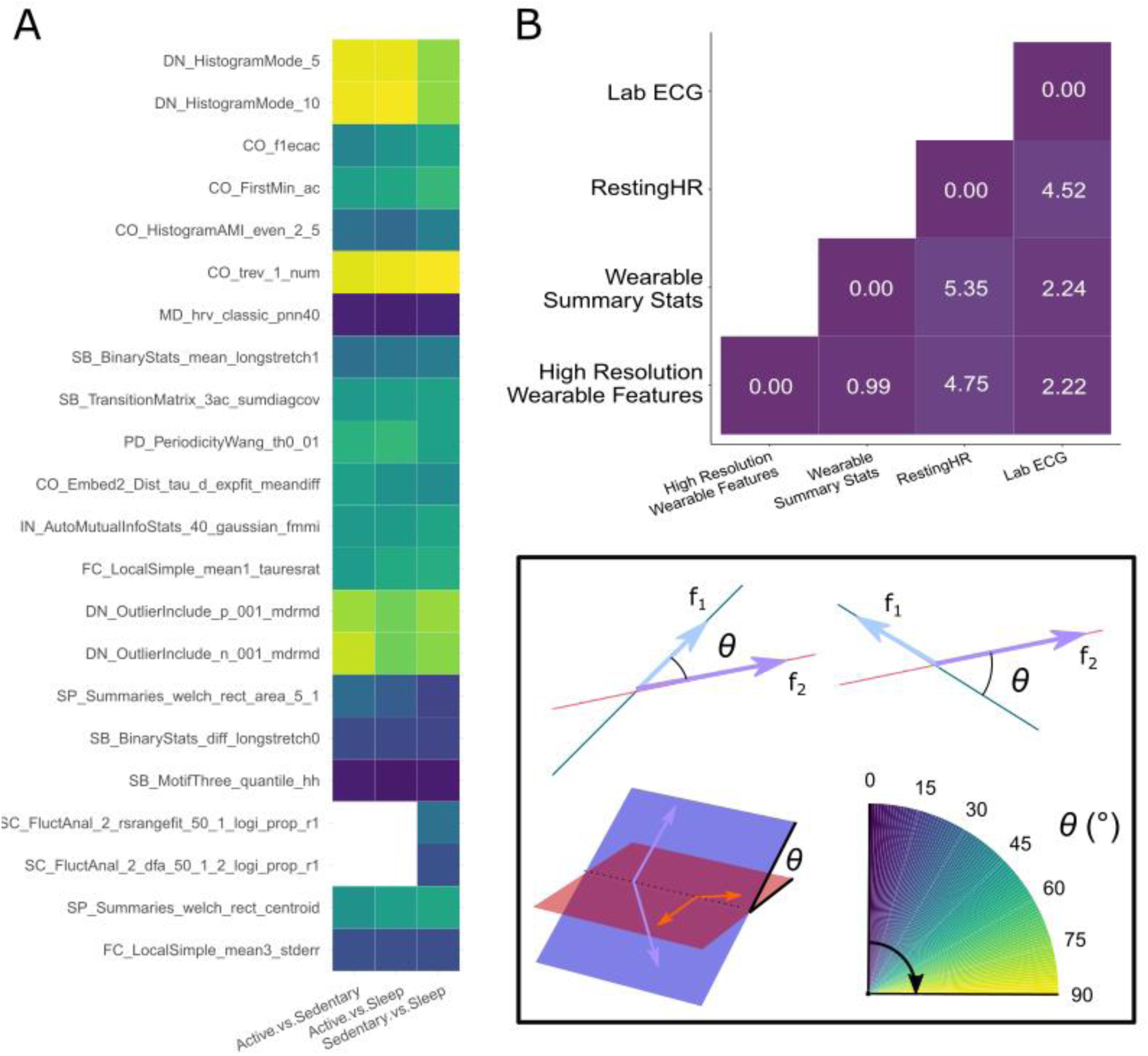
Relationship Between Different Heart-Rate Features. Angular distance, θ (°), was used to assess similarity between features. (A) Angles between high resolution feature from the three different activity periods. White colour is used where the angle was undefined. (B) Subspace angles between the three wearable feature sets and the laboratory ECG measurements.

Finally, we evaluated inter-relations between the different feature sets, namely the high resolution wearable features, RestingHR, wearable summary statistics, and the clinical gold standard ECG features (PR, QRS, QT, ECG.HR and QTc). Specifically, to evaluate the degree of overlap between information from the different feature sets, we computed the subspace angle between the feature matrices of interest (Figure 3B). For this pairwise computation between feature sets, we selected a common set of 315 subjects who had valid (i.e., non-null) records for all the features under consideration. Intuitively, two feature sets with independent information content would be orthogonal (90°) to each other, whereas two collinear feature sets would have subspace angle of 0°; the smaller the subspace angles between two feature sets, the more overlap in the information content. Overall, the different heart rate features exhibit substantive overlap (θ < 6°). However, the high resolution and summary statistic features derived from the wearables are most distinct from RestingHR (θ = 4.75° and 5.35° respectively). Further, the clinical gold standard features obtained via laboratory ECG measurements are most distinct from wearable-based RestingHR (θ = 4.52°), but have good overlap with both the wearable-derived high-resolution features and wearable summary statistics (θ = 2.22° and 2.24° respectively). These findings suggest that clinically informative features could be obtained with consumer devices in home or community settings.

### Predictive Value of Different Wearable-Derived Feature Sets for Clinical Targets

Having gained some intuition about the information contained within the wearable-derived feature sets, we considered their predictive value for the clinical markers of cardiometabolic disease risk. Specifically, we trained random forest models to use the different wearable-derived feature sets for classification of each of the four cardiometabolic disease risk targets. We performed two sets of comparative analyses to evaluate predictive value of the wearable-derived feature sets for classification of different cardiometabolic disease risk targets, and detail findings below.

#### Association with Clinical Risk Markers

First, we compared the out-of-bag performance of models trained using different feature sets for each clinical risk marker target (Figure 4). For each target, the best performance model was based on one of the high resolution wearable feature sets (HighRes.ActiveSeg, HighRes.SedenSeg or HighRes.SleepSeg). This finding highlights the predictive value of the high resolution information within wearable-derived heart rate time series recordings.

**Figure 4.**
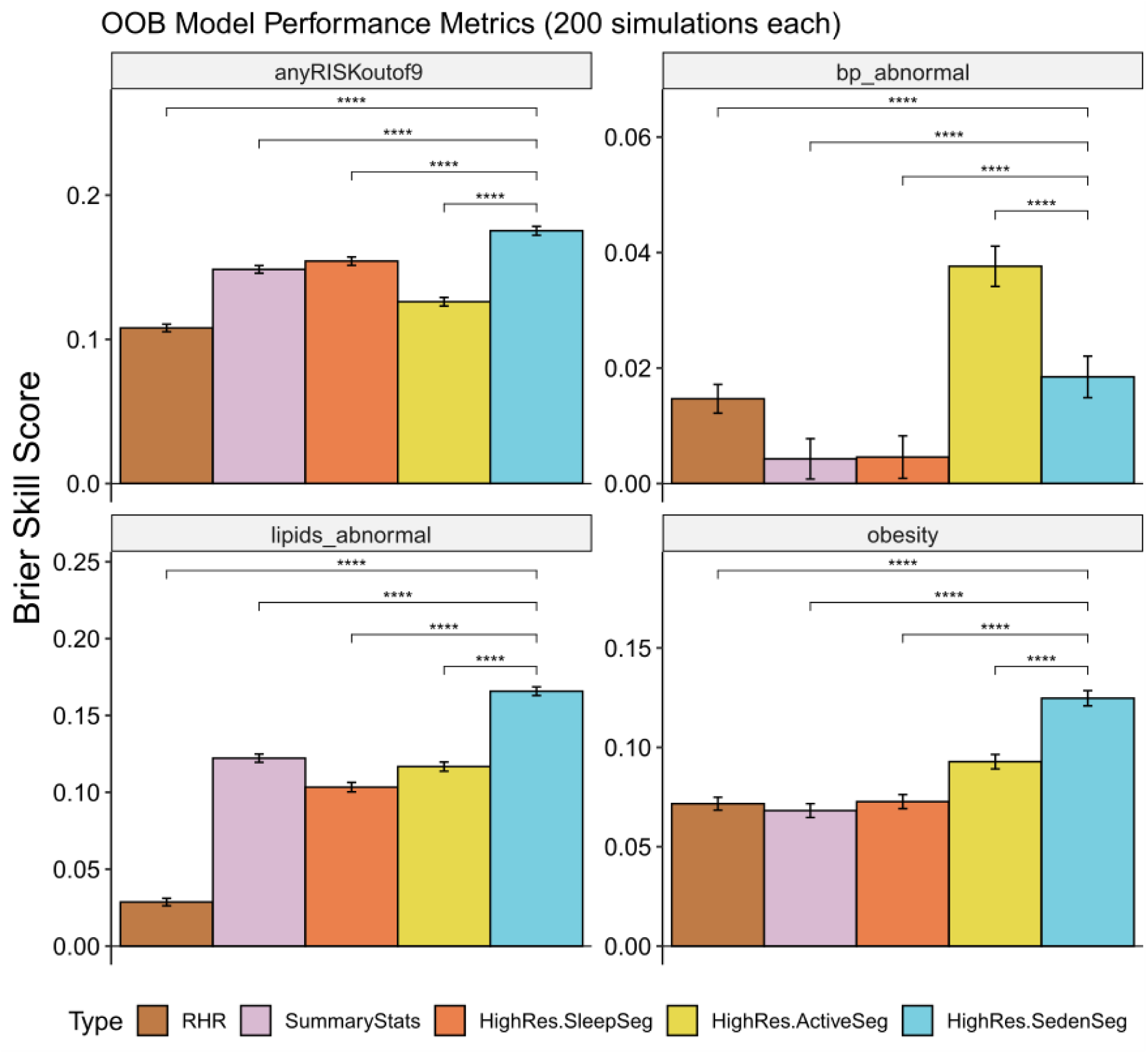
Model Performance on Cardiometabolic Risk Targets. Model performance for each of the five model types computed for the four targets. A higher Brier Skill Score indicates better performing model. The baseline model on which the Brier Skill Score is based on only has age and gender as features; feature sets used in each model type are detailed in Table 1. Comparisons of Skill Scores from the different models against those from the HighRes.SedenSeg model (using *t*-tests) indicate significant differences, with BH-adjusted *P* values all being <.001 (represented by ‘****’).

Second, we observe that heart rate dynamics extracted from different activity level segments have differential predictive potential for the various targets, evidenced by the statistically significant differences between Brier skill scores of the HighRes.ActiveSeg, HighRes.SedenSeg and HighRes.SleepSeg models (Figure 4). Of the three model types, HighRes.SedenSeg performs best for lipid abnormalities, obesity and anyRISKoutof9; while HighRes.ActiveSeg performs best for blood pressure abnormalities.

Third, to comparatively evaluate contributions from individual wearable-derived features, we trained models that utilize all features available to predict each cardiometabolic disease risk target, and ranked the variable importance in each case. Figure 5 shows the variable importance plots, averaged across 200 independent simulations, based on the combined set of top ten ranking features for each of the four cardiometabolic disease risk targets. It is immediately clear that different features drive the performance of the models for each of the four targets. For instance, age and gender are the top two drivers of model performance for the anyRISKoutof9 target, but are not even within the top ten for both lipids_abnormal and obesity. Similarly, the set of features that primarily drives the performance for lipids_abnormal includes many that are detrimental to performance for the anyRISKoutof9 target. Further, our findings show that heart rate dynamics from different activity states contain distinct information on cardiometabolic disease risk. For example, the DN_HistogramMode_5 feature from the sedentary and active segments is important for predicting the cardiometabolic disease risk markers (Figure 5), but the DN_HistogramMode_5 feature from the sleep segment is not.

**Figure 5.**
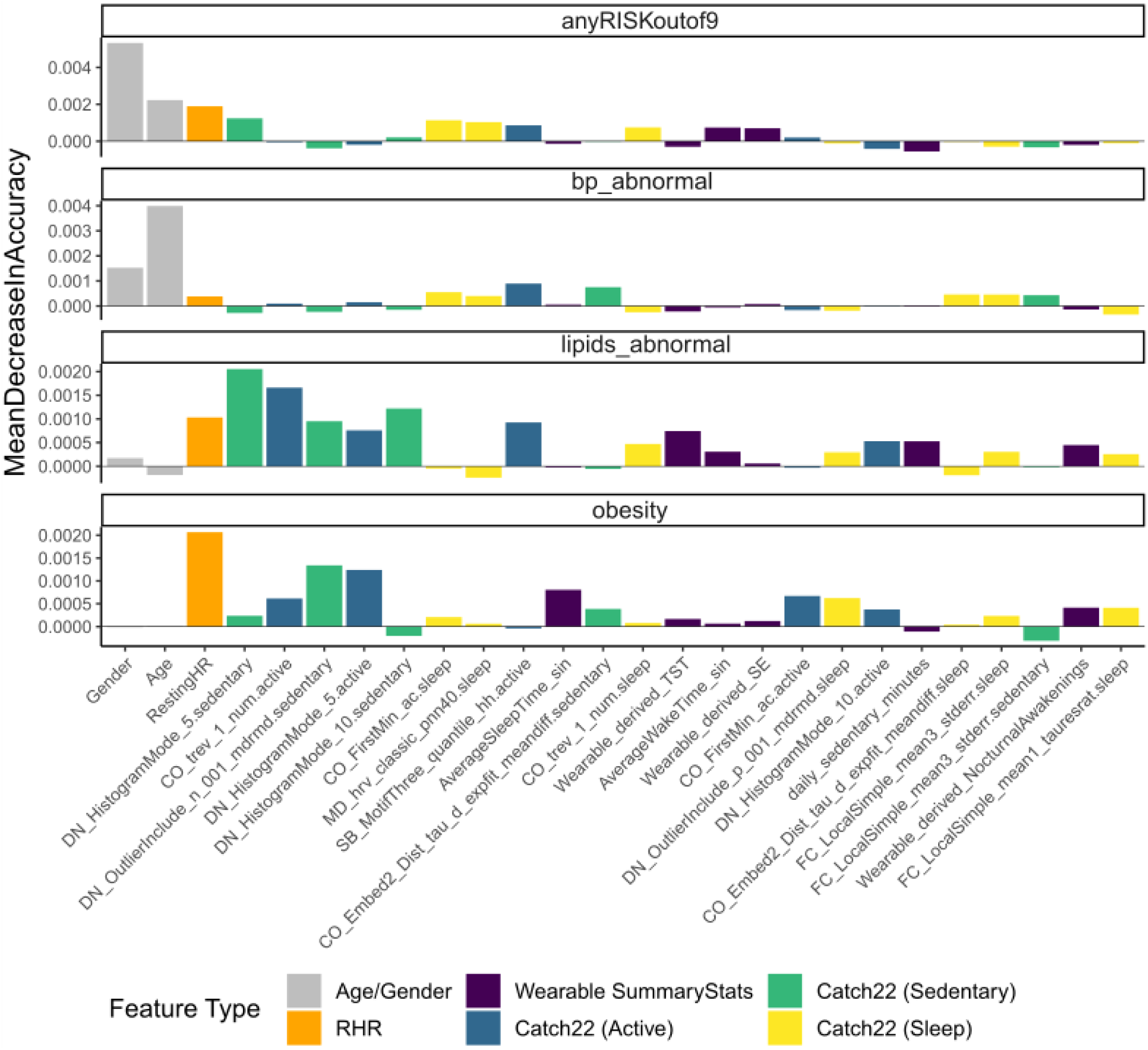
Random Forest Variable Importance. The variable importance of each feature for prediction of the four cardiometabolic disease risk targets. We averaged each importance value across 200 simulations, and used the results to rank the top ten features to retain for each cardiometabolic disease risk target. This resulted in a total of 26 features across all four targets, as shown.

Fourth, we observe that the top ten features for each of the four targets included features from all six feature types (age/gender, RHR, wearable summary statistics, and the three sets of high resolution features from Table 1). This suggests that risk prediction models using wearable-derived features may not exclusively rely on only one of the different feature sets, or any one feature drawn from these feature sets for that matter. Rather, a collection of different wearable-derived high resolution heart rate features from distinct activity states is essential to accurately predict a multiplicity of cardiometabolic disease risk targets.

#### Illustrative Case Studies for Clinical Events

Finally, we examined relations between the most predictive wearable-derived feature set (i.e., Catch22 (Sedentary)) and actualized clinical events. Of the 692 subjects in our study, we filtered those who actualized clinical events for cardiometabolic conditions during the longitudinal monitoring period after initial data collection. Through this process, we identified a total of 11 subjects who developed relevant cardiometabolic conditions: cardiovascular disease (CVD; five subjects), dyslipidemia (four subjects) and hypertension (two subjects). None of the subjects had actualized obesity related events. Of these 11 subjects, we further selected those subjects who did not overlap with the set of 321 subjects whose data were used to train predictive models for clinical risk markers. This yielded a set of four subjects: one subject had ICD codes for all three conditions, two subjects only had codes for dyslipidemia, and one subject only had codes for cardiovascular disease. Amongst these four, we selected the two subjects with cardiovascular disease for further profiling: one with CVD, dyslipidemia and hypertension (Subject A) and one with CVD only (Subject B).

For these two subjects, we visualized the 22 wearable-derived high resolution features using two clockplots (see SI Table 3 for the feature names corresponding to the numeric IDs). First, for a given subject, we plotted the percentile value of each feature in relation to the associated distribution in the training dataset (321 subjects). We term this the feature value percentile clockplot. Any feature exhibiting extreme percentile values stands out in relation to its typical distribution across individuals in our training set. Second, for each feature, we considered a cluster of the 10 training set members that are most similar (based on feature value proximity) to the subject of interest; and plotted the percentage of the cluster members who have positive anyRiskoutof9 labels. The percentage of positive labels in this cluster suggests the degree to which this *individual* feature value is indicative of risk. We term this the positive label proportion clockplot.

We present the profiles and some illustrative findings for subject A and B in Figures 6A and 6B, respectively.

**Figure 6.**
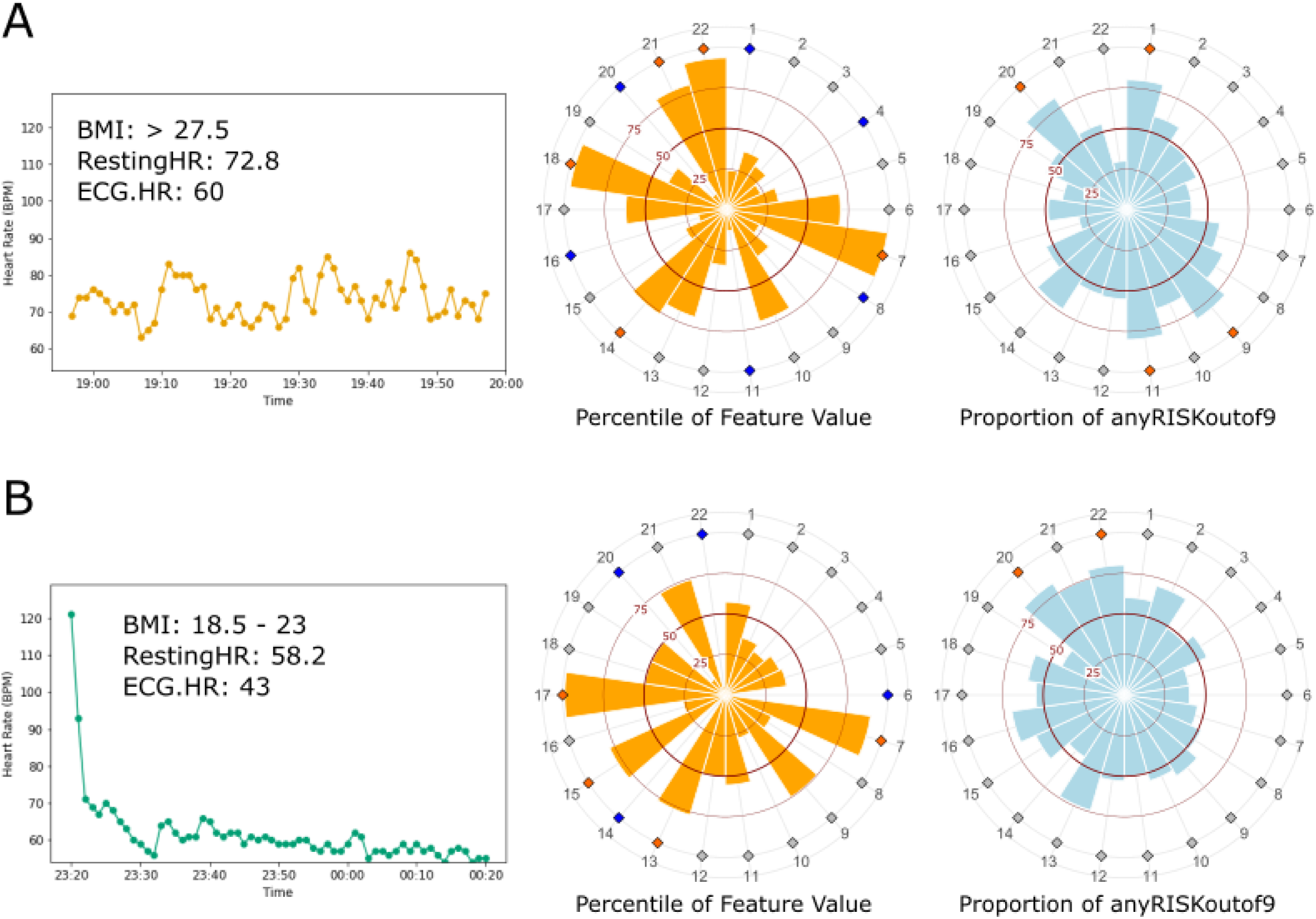
Illustrative Profiles of Two Subjects with Actualized Cardiometabolic Disease events. Subjects A and B with associated ranges of BMI, RestingHR and laboratory ECG heart rate measurements. The heart rate time series corresponds to the 1-hour sedentary period used to generate the high resolution features. The two clockplots present the 22 high-resolution features obtained for the subject; the left plot depicts the percentile value of each feature in relation to the associated training set distribution, while the right plot shows the local likelihood that a given feature value would be seen in individuals who are positive for anyRISKoutof9. For both clockplots, red diamonds are used to indicate features with values exceeding the 75^th^ percentile or 75% proportion, and blue diamonds for features with values below the 25^th^ percentile or 25% proportion. The complete list of feature names corresponding to the numerical IDs in the clockplots can be found in SI Table 3.

Subject A was a 51-55 year-old male who was assessed at the start of the study to have very high BMI, higher than average wearable-derived resting heart rate of 72.8 bpm, and abnormal blood pressure and sugar levels. Amongst the wearable-derived high resolution features from the sedentary period, features 1, 11 and 20 (DN_HistogramMode_5, CO_Embed2_Dist_tau_d_expfit_meandiff and SC_FluctAnal_2_dfa_50_1_2_logi_prop_r1 respectively) stand out for having extreme percentile values (feature value percentile clockplot) that are more typical of training subjects with positive labels for anyRISKoutof9 (positive label proportion clockplot). We note that the feature 1 and 11 are the most important high resolution features in the corresponding HighRes.SedenSeg model for anyRISKoutof9 (See SI-7).

In contrast, subject B was a 56-60 year-old male with a seemingly healthier profile, having lower BMI and lower than average resting heart rate of 58.2 bpm. Accordingly, subject B only had abnormal lipids levels and no other risk markers at the start of the study. However, the high resolution features characterizing this subject depict a richer picture. Eight out of 22 features exhibited values either below the 25^th^ percentile or above the 75^th^ percentile of their corresponding training set distributions (feature value percentile clockplot). Of these, features 6, 14 and 17 (CO_trev_1_num, DN_OutlierInclude_p_001_mdrmd and SB_BinaryStats_diff_longstretch0, respectively) had values at the 5.9^th^, 0^th^ and 98.8^th^ percentile of their corresponding training set distributions, respectively. We note that features 6, 14 and 17 are amongst the top ten most important features for the corresponding HighRes.SedenSeg model for anyRISKoutof9 (See SI-7).

As neither Subject A nor Subject B were part of the training set used to develop the models, the above observations are consistent with the hypothesis that there are true associations between some of the high resolution features and the cardiometabolic disease risk targets. Although extreme feature values, in and of themselves, may not always determine the eventuality of a cardiovascular disease event, the above comparisons illustrate the discriminative potential of the high resolution wearable-derived heart rate features over and above evident BMI and heart rate measures. These illustrative case studies also highlight that the specific subset of wearable features taking on extreme values may be different for different individuals. This suggests the need for a diverse set of high resolution heart rate features, and a model that allows interactions between these features, in order to better explain potential risks. We finally note that the lifestyle and genomic markers for these subjects, detailed in SI-8, are largely similar and differences that do exist may reflect in the heart rate and BMI profiles we considered above. The above observations collectively suggest that the wearable-derived high resolution heart rate features could contain additional physiological information beyond typical self-reported health and wellness metrics and/or common summary statistics used to assess cardiometabolic disease risk.

### Associations between Wearable-Derived Features, Genetic and Lifestyle Markers

To further interpret the information contained within the wearable-derived features, we sought to understand how they relate to genetic predispositions for cardiometabolic disease, lifestyle habits and health perceptions. In particular, we focused these analyses on the commonly used RestingHR feature, the wearable-derived summary statistics feature set, as well as the high resolution feature set with most predictive value for all the clinical risk markers combined, i.e., Catch22 (Sedentary) (see Figure 4: anyRISKoutof9 panel); the corresponding models were RHR, SummaryStats and HighRes.SedenSeg respectively (Table 1).

#### Associations with Genetic Risk Scores

We examined the degree of information overlap between the different wearable-derived features (Table 1) and the genetic risk for cardiometabolic conditions. For each pairing between the three wearable-derived feature sets and the three genetic risk targets, we trained random forest models, and used the Brier Skill Scores of the different feature sets (against a baseline model that only included gender as a covariate) as indirect measures of strength of the associations.

The results are in Figure 7. For each of the three abnormality types, we observe that the high resolution wearable features were more strongly associated with genetic risk levels than RestingHR. Further, for genetic predisposition to lipid abnormalities and obesity, the high resolution wearable-derived heart rate feature set had stronger associations than the summary statistics feature set. However, for genetic predisposition to high blood pressure, the summary statistics features had a slightly stronger association than the high resolution wearable-derived features (yet with small 0.025 margin in Brier Skill Score between HighRes.SedenSeg and SummaryStats models). We highlight that these trends are relatively insensitive to the polygenic risk score threshold used for defining high vs. low genetic risk (SI-6). These results suggest that the wearable recordings may capture physiological dynamics related to genetic risk for cardiometabolic disease.

**Figure 7.**
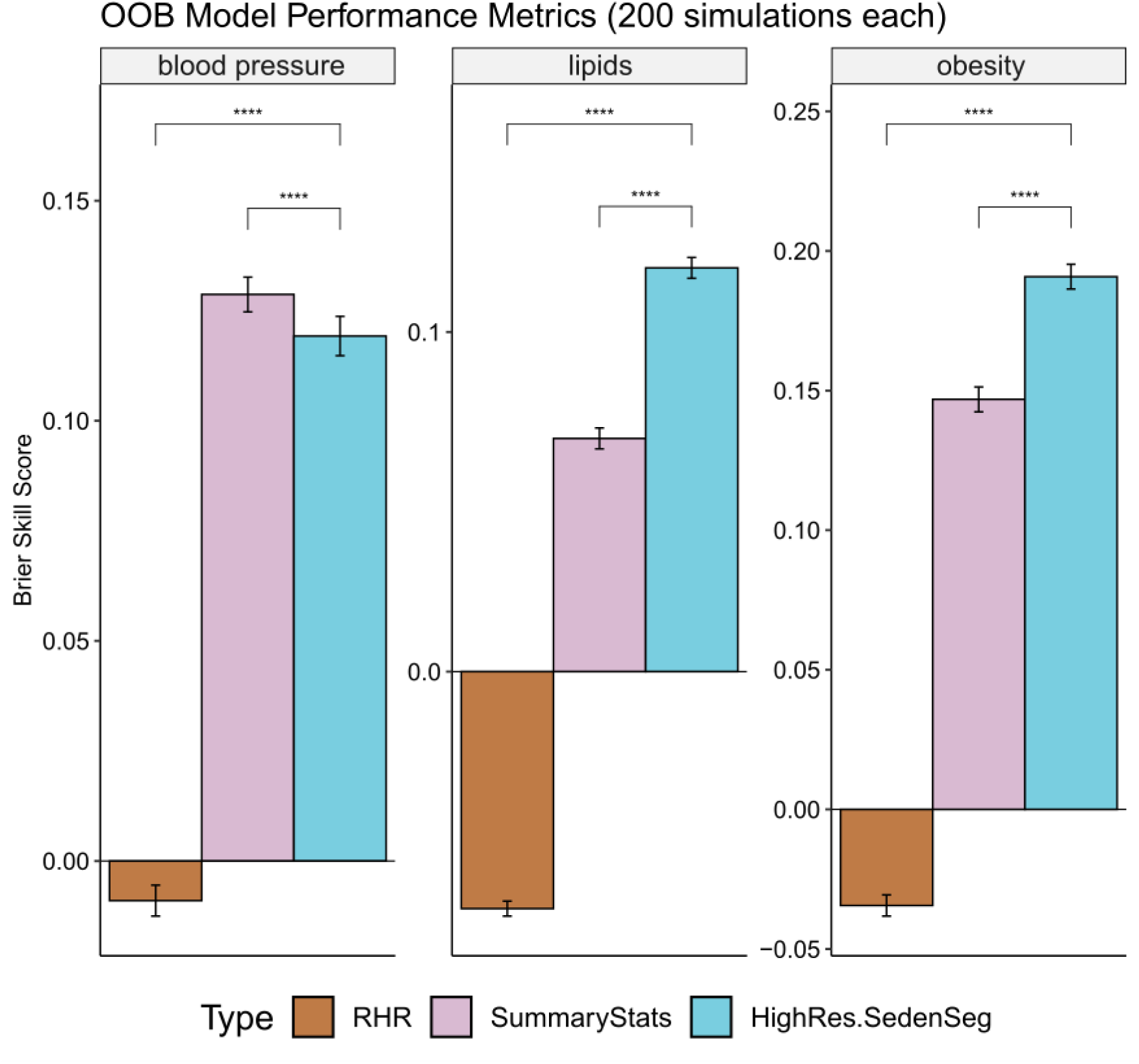
Degree of Association with Genetic Risk Targets. Out-of-bag (OOB) performance for each of the five model types computed for the three targets. A higher Brier Skill Score indicates better performing model; negative scores indicate that the model performs worse than the comparative baseline model. The baseline model used for the Brier Skill Score computations has gender as the only covariate. RHR: Baseline + RestingHR; SummaryStats: Baseline + Summary Statistics; HighRes.SedenSeg: Baseline + Catch22 (Sedentary).

#### Association with Lifestyle Habits and Health Perceptions

Next, we studied overlap between the different wearable-derived feature sets, and general lifestyle habits (LH) and health perceptions (HP). For each combination of the three wearable-derived feature sets and the seven LP and HP targets, we trained random forest models, and used the Ranked Probability Skill Scores of the different feature sets (against a baseline model with gender and age as covariates) as indirect measures of strength of the associations.

We first examine the results for health perceptions (Figure 8A-D). For all these cases, the wearable-derived summary statistics and high resolution features have stronger associations than resting heart rate alone. In particular, the wearable-derived summary statistics are highly correlated with states pertaining to stress, anxiety and depression. This is intuitive as these states affect many aspects of a subject’s activity and sleep-wake patterns. Intriguingly, however, we find that the wearable-derived high resolution heart rate features are highly associated with pain and discomfort, as well as with the overall perceived health state. This suggests that heart rate dynamics contain indicative information on overall pain levels, and health and wellness perceptions. Hence, wearable heart rate recordings may serve as objective measures for these often very subjective and difficult to assess perceptions.

**Figure 8.**
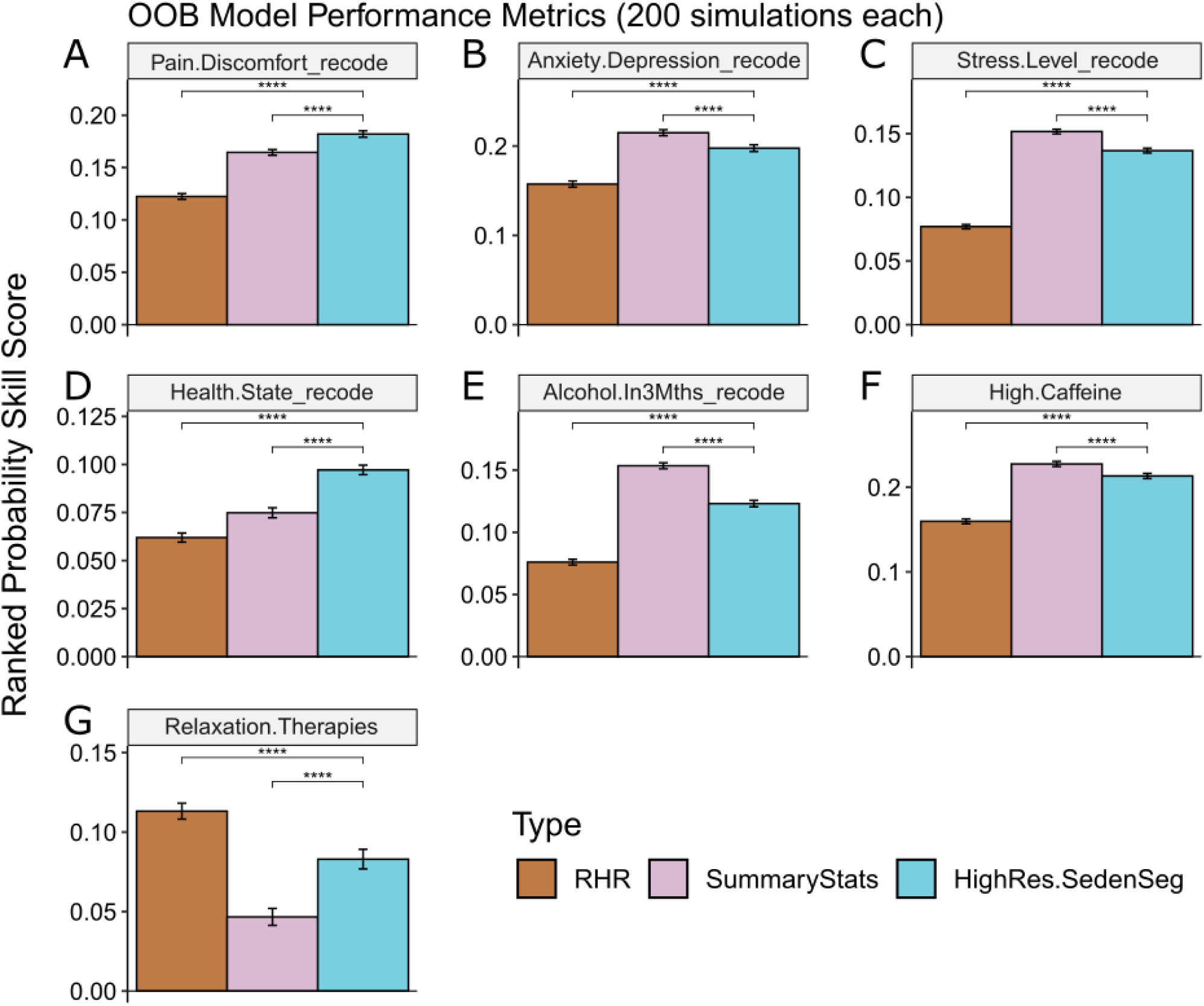
Model Performance on Lifestyle Habits and Health Perception Targets. Out-of-bag (OOB) performance for each of the three model types computed for the seven targets. A higher Brier Skill Score indicates better performing model. The baseline model used for computing the Ranked Probability Skill Score uses age and gender as covariates. RHR: Baseline + RestingHR; SummaryStats: Baseline + Summary Statistics; HighRes.SedenSeg: Baseline + Catch22 (Sedentary).

Next, we turn to the results for lifestyle habits (Figure 8E-G). We observe that RestingHR is most strongly associated with engaging in relaxation therapies. This suggests that the overall (average) heart rate may have more information on relaxation than higher resolution heart rate dynamics. On the other hand, for the consumption patterns of alcohol and caffeinated drinks, the higher resolution feature sets have stronger associations than RestingHR alone. In particular, wearable-derived summary statistics are most associated with alcohol consumption habits while wearable-derived high resolution features are most associated with caffeine consumption. Contrary to expectations, resting heart rate has weaker associations with caffeine consumption than higher resolution features derived from sedentary segments. This suggests that dynamics (e.g., irregularity or “spikiness”) of the heart rate time series even while a subject is sedentary may be associated with caffeine consumption habits. While the size of our data and data collection process do not enable assessments of causality of the above relationships, the above results provide interesting insights into how physiological dynamics that manifest in wearable recordings may be correlated with lifestyle habits.

## Discussion

Consumer wearables enable recording of rich high resolution physiological dynamics in free-living conditions, but how these data relate to health and disease is not fully understood. We introduced a principled framework to derive high resolution heart rate features from consumer wearable recordings. We applied our approach on a dataset containing multidimensional cardiometabolic health parameters from healthy volunteers, and demonstrated the utility of high resolution wearable features in understanding cardiometabolic disease risk. Our results highlight the additional value of these high resolution features over typical summary statistics, and show that wearable data recorded on an ongoing basis are associated with genetic predispositions and lifestyle habits alike. Therefore, we posit that high resolution digital phenotypes from consumer wearables could find potential use in longitudinal monitoring of cardiometabolic health.

Our framework is customized to address key challenges in mining wearable data recorded in free-living conditions. Unlike clean data from controlled experimental settings, real-world wearable recordings tend to be irregular, contain missing stretches [29], lack clean context annotations, and have variable lengths. As such, analyses based on naïve application of general-purpose time series feature extraction methods [37,40,57] may not have ecological validity [58]. To address this gap and derive meaningful physiological dynamics from wearable time series recordings, our feature extraction framework standardizes handling of data irregularities, and encodes contextual information about underlying activity level and physiological state (Figure 1-3). This conceptual framework, although demonstrated here with the Catch22 method [40], is agnostic to choice of time series featurization methods [37,38]. Further, in contrast to black-box feature learning methods based on large labelled datasets [31], our approach yields more interpretable time series features with smaller unlabelled datasets.

Our framework enables many possibilities for gaining new insights with wearable recordings. To illustrate this, we analysed multimodal wearable, genomic, lifestyle and clinical data from healthy volunteers and highlight two of these possibilities.

First, our results reveal new relations between high resolution heart rate dynamics from wearables and risk of cardiometabolic disease. Most previous studies correlate clinically obtained measures of heart rate dynamics, such as heart rate variability, exercise capacity, and heart rate recovery, with disease risk or outcomes [59–61]. In contrast, our results reveal that heart rate dynamics recorded by consumer wearables, when processed appropriately, are also predictive of cardiometabolic disease risk (Figures 4 and 6). Further, we find that heart rate dynamics from different activity states contain distinct information about specific cardiometabolic conditions (Figures 2-5). For example, heart rate patterns from sedentary states are more related to lipid abnormalities and obesity, whereas those from active states may be more related to blood pressure abnormalities (Figure 4). These findings highlight the value-add of assessing physiology in free-living activity states (beyond controlled clinical settings) for disease risk monitoring [62].

Second, our work provides new perspectives on interfaces between wearable recordings, genetic predispositions and lifestyle factors in cardiometabolic disease. Although there has been longstanding interest in probing gene-lifestyle interactions and their additive effects on cardiovascular disease [63–65], such studies have had limited visibility into physiology in free-living conditions. We found surprising connections (Figure 7) between our wearable-derived features and genetic predispositions for cardiometabolic disease. As these associations did not appear to depend on the presence or absence of manifest clinical risk markers, we posit that high resolution phenotypes from wearables may capture subtle subclinical physiological changes stemming from latent predispositions to disease. Moreover, while wearables are known to capture intimate details on daily life patterns [66,67], our results suggest that high resolution features in wearable records could serve as objective indicators of subjective perceptions of stress, anxiety, pain and overall health state (Figure 8). Collectively, these findings suggest that high resolution digital phenotypes from wearables could provide a means to passively but objectively assess physiological changes relating to a host of nature and nurture factors.

While the uniquely multimodal nature of our data enables us to uncover many novel insights on high-resolution wearable phenotypes, limitations of dataset size and cohort design present some challenges. For instance, it was infeasible to conduct full-scale gene-environment interaction studies [68–70]; analyse relevant lifestyle factors such as smoking (as only 9 smokers had wearable records of sufficient durations); or train state-of-the-art machine learning models with large feature sets. Further, as the study exclusively enrolled healthy subjects, only 11 subjects subsequently presented with actualized cardiometabolic events in the longitudinal monitoring period, limiting our clinical outcome evaluations. Future work based on larger cohorts [71] with more longitudinal follow-up could address some of these limitations. Such efforts would also enable cross-cohort validation of our current findings; expansion of our findings to even higher resolution digital phenotypes that can be extracted from recordings with newer generations of wearable devices [72,73]; and targeted evaluations of value for precision screening, health monitoring and disease management applications.

## Data Availability

All data produced in the present study are available upon reasonable request to the authors

## Acknowledgements

This research was supported by funding and infrastructure from the Singapore National Precision Medicine programme (IAF-PP H17/01/a0/007) and the Institute for Infocomm Research, A*STAR. The data acquisition was supported in part by funding from SingHealth, Duke-NUS Medical School, National Heart Centre Singapore (NHCS), Singapore National Medical Research Council (NMRC/STaR/0011/2012, NMRC/STaR/0026/2015), Lee Foundation, and Tanoto Foundation.

We would like to thank all volunteers for their participation in this study. We acknowledge valuable data collection assistance from NHCS and SingHEART Clinical Research Coordinators; and resources from the National Supercomputing Centre, Singapore (https://www.nscc.sg) for code development on a polygenic score computation pipeline. We also thank Marie Loh from the Nanyang Technological University, Singapore and Xueling Sim from the National University of Singapore for meaningful discussions on polygenic risk scores.

J.Z. was affiliated with the Institute of Infocomm Research at the time of his contribution to this work and is currently affiliated with Diagnostics Development Hub (DxD Hub) at the Agency for Science Technology and Research (A*STAR).

## Abbreviations

PGS: Polygenic Score
ECG: Electrocardiogram
ICD-10: International Classification of Diseases, 10^th^ Revision

