## SupplementaryInformation for "High-Resolution Digital Phenotypes from Consumer Wearables Enhance Prediction of Cardiometabolic Risk Markers"

**Author Affiliations**

### Supplementary Information

#### SI Table 1: Data Distribution

| | Female (*n* = 370; 53.5%) | | | Male (*n* = 322; 46.5%) | | |
|---|---|---|---|---|---|---|
| | **Mean** | **Std Dev** | **# NA** | **Mean** | **Std Dev** | **# NA** |
| **Age, years** | 45.47 | 11.71 | 0 | 44.46 | 13.29 | 0 |
| **BMI, kg/m2** | 22.87 | 3.94 | 0 | 24.33 | 3.39 | 0 |
| **WC, cm** | 78.91 | 10.98 | 0 | 86.96 | 9.86 | 0 |
| **SBP, mmHg** | 122.5 | 17.74 | 0 | 132.2 | 14.96 | 0 |
| **DBP, mmHg** | 73.38 | 12.80 | 0 | 82.18 | 10.97 | 1 |
| **RestingHR, (Fitbit, bpm)** | 70.66 | 6.55 | 0 | 69.37 | 6.59 | 0 |
| **ECG_HR, bpm** | 64.46 | 9.17 | 10 | 63.67 | 9.87 | 12 |
| **Total Cholesterol, mmol/l** | 5.345 | 0.94 | 6 | 5.332 | 0.97 | 5 |
| **LDL, mmol/l** | 3.316 | 0.81 | 7 | 3.399 | 0.89 | 6 |
| **HDL, mmol/l** | 1.586 | 0.32 | 6 | 1.357 | 0.30 | 5 |
| **TGs, mmol/l** | 0.9858 | 0.51 | 6 | 1.3 | 0.76 | 5 |
| **Glucose, mmol/L** | 5.172 | 0.49 | 8 | 5.358 | 0.71 | 5 |
| **Average Daily Step Count** | 11585.6 | 4898.24 | 10 | 12101.06 | 4435.12 | 10 |
| **Average Daily Sedentary Minutes** | 633.5 | 96.48 | 102 | 656.5 | 95.58 | 88 |
| **Average Daily Sleep Hours** | 395.9 | 61.18 | 102 | 374.5 | 65.15 | 88 |

#### SI Table 2: Wearable Data Summary Statistics

| Feature Set Type | Features | Description |
|---|---|---|
| Summary Statistics for Mean Daily Physical Activity Durations | Wearable_derived_TST | Average wearable-derived total sleep time |
| | daily_sedentary_minutes | Average minutes/day spent in sedentary period |
| | daily_active_minutes | Average minutes/day spent in active period |
| Summary Statistics from Device Logs | Wearable_derived_Nocturnal Awakenings_minutes | Average daily minutes of nocturnal awakenings |
| | Wearable_derived_Nocturnal Awakenings | Average number of nocturnal awakenings |
| | Wearable_derived_SE | Wearable-derived sleep efficiency score (from Fitbit) |
| Average Wake and Sleep Times | AverageWakeTime_sin | Mean waking time, sine transformed |
| | AverageWakeTime_cos | Mean waking time, cosine transformed |
| | AverageSleepTime_sin | Mean bedtime, sine transformed |
| | AverageSleepTime_cos | Mean bedtime, cosine transformed |

Sleep efficiency used to be a score that could be retrieved by the Fitbit API. The formula has never been published by Fitbit, although a comparison of actual sleep records against the retrieved scores indicates that it is defined as:

$$\text{Sleep Efficiency} = \frac{minutesAsleep}{minutesAsleep + minutesAwake} \times 100$$

### SI Table 3: Description of Catch22 Features

**NB: Descriptions are reproduced from Table 1 of Lubba et al**

| ID | Feature Name | Description |
|---|---|---|
| 1 | DN_HistogramMode_5 | Mode of z-scored distribution (5-bin histogram) |
| 2 | DN_HistogramMode_10 | Mode of z-scored distribution (10-bin histogram) |
| 3 | CO_f1ecac | First 1/e crossing of autocorrelation function |
| 4 | CO_FirstMin_ac | First minimum of autocorrelation function |
| 5 | CO_HistogramAMI_even_2_5 | Automutual information, $m=2,\tau=5$ |
| 6 | CO_trev_1_num | Time-reversibility statistic, ⟨(xt+1−xt)3⟩t |
| 7 | MD_hrv_classic_pnn40 | Proportion of successive differences exceeding $0.04\sigma$ |
| 8 | SB_BinaryStats_mean_longstretch1 | Longest period of consecutive values above the mean |
| 9 | SB_TransitionMatrix_3ac_sumdiagcov | Trace of covariance of transition matrix between symbols in 3-letter alphabet |
| 10 | PD_PeriodicityWang_th0_01 | Periodicity measure |
| 11 | CO_Embed2_Dist_tau_d_expfit_meandiff | Exponential fit to successive distances in 2D embedding space |
| 12 | IN_AutoMutualInfoStats_40_gaussian_fmmi | First minimum of the automutual information function |
| 13 | FC_LocalSimple_mean1_tauresrat | Change in correlation length after iterative differencing |
| 14 | DN_OutlierInclude_p_001_mdrmd | Time intervals between successive extreme events above the mean |
| 15 | DN_OutlierInclude_n_001_mdrmd | Time intervals between successive extreme events below the mean |
| 16 | SP_Summaries_welch_rect_area_5_1 | Total power in lowest fifth of frequencies in the Fourier power spectrum |
| 17 | SB_BinaryStats_diff_longstretch0 | Longest period of successive incremental decreases |
| 18 | SB_MotifThree_quantile_hh | Shannon entropy of two successive letters in equiprobable 3-letter symbolization |
| 19 | SC_FluctAnal_2_rsrangefit_50_1_logi_prop_r1 | Proportion of slower timescale fluctuations that scale with linearly rescaled range fits |
| 20 | SC_FluctAnal_2_dfa_50_1_2_logi_prop_r1 | Proportion of slower timescale fluctuations that scale with DFA (50% sampling) |
| 21 | SP_Summaries_welch_rect_centroid | Centroid of the Fourier power spectrum |
| 22 | FC_LocalSimple_mean3_stderr | Mean error from a rolling 3-sample mean forecasting |

#### SI Table 4: Description of ICD codes used

##### Cardiovascular Disease

| | ICD 10 Code | Description |
|---|---|---|
| 1 | I200 | Unstable angina |
| 2 | I208 | Other forms of angina pectoris |
| 3 | I211 | Acute transmural myocardial infarction of inferior wall |
| 4 | I214 | Acute subendocardial myocardial infarction |
| 5 | I2511 | Atherosclerotic heart disease, of native coronary artery |
| 6 | I258 | Other forms of chronic ischaemic heart disease |
| 7 | I259 | Chronic ischaemic heart disease, unspecified |
| 8 | I420 | Dilated cardiomyopathy |
| 9 | I440 | Atrioventricular block, first degree |
| 10 | I447 | Left bundle-branch block, unspecified |
| 11 | I451 | Other and unspecified right bundle-branch block |
| 12 | I458 | Other specified conduction disorders |
| 13 | I471 | Supraventricular tachycardia |
| 14 | I48 | Atrial fibrillation and flutter |
| 15 | I493 | Ventricular premature depolarisation |
| 16 | I495 | Sick sinus syndrome |
| 17 | I498 | Other specified cardiac arrhythmias |
| 18 | R000 | Tachycardia, unspecified |
| 19 | R001 | Bradycardia, unspecified |

##### Dyslipidemia

| | ICD 10 Code | Description |
|---|---|---|
| 1 | E780 | Pure hypercholesterolemia |
| 2 | E781 | Pure hyperglyceridemia |
| 3 | E782 | Mixed hyperlipidemia |
| 4 | E783 | Hyperchylomicronemia |
| 5 | E784 | Other hyperlipidemia |
| 6 | E785 | Hyperlipidemia, unspecified |
| 7 | E786 | Lipoprotein deficiency |

##### Hypertension

| | ICD 10 Code | Description |
|---|---|---|
| 1 | I10 | Essential (primary) hypertension |
| 2 | I11 | Hypertensive heart disease |
| 3 | I12 | Hypertensive chronic kidney disease |
| 4 | I13 | Hypertensive heart and chronic kidney disease |

##### Obesity

| | ICD 10 Code | Description |
|---|---|---|
| 1 | E668 | Other obesity |
| 2 | E669 | Obesity, unspecified |

### SI-1: Determination of Time Series Segment Length for Catch22 Features

For each individual, we obtained the longest continuous heart rate time series in active, sedentary, and sleep periods. The median lengths of the time series are 31mins, 1h 45mins, and 7h 45mins respectively (1). To determine what the ideal time length should be for generating catch22 features in each of those three periods, we ran a series of experiments in the following manner. First, we computed catch22 on sliding windows of varying lengths for each of the three activity periods (active: [10min, 15min, 20min, 25min, 30min], sedentary: [10min, 20min, 30min, 1h], sleep: [10min, 20min, 30min, 1h, 3h, 5h]). We then calculated the coefficient of variation (CV) of each feature for each individual, and averaged it across all eligible individuals. For active, 20mins yielded the most stable results. For sedentary, 1h was the most stable. And lastly, for sleep, 5h was the most stable. (2) We thus used the first 20 mins, 1h, and 5h of the longest continuous heart rate time series in active, sedentary and sleep periods respectively, to generate three sets of catch22 features for each individual.

#### Summary Statistics

##### Summary stats of longest continuous TS in active period

| | |
|---|---|
| Count | 642 |
| Mean | 0 days 00:38:00.654205607 |
| Std | 0 days 00:22:26.685840823 |
| Min | 0 days 00:07:00 |
| 25% | 0 days 00:22:00 |
| 50% | 0 days 00:31:00 |
| 75% | 0 days 00:47:00 |
| Max | 0 days 02:39:00 |

##### Summary stats of longest continuous TS in sedentary period

| | |
|---|---|
| Count | 629 |
| Mean | 0 days 03:07:24.133545310 |
| Std | 0 days 02:48:02.781473861 |
| Min | 0 days 00:16:00 |
| 25% | 0 days 01:10:00 |
| 50% | 0 days 01:45:00 |
| 75% | 0 days 04:38:00 |
| Max | 0 days 19:37:00 |

##### Summary stats of longest continuous TS in sleep period

| | |
|---|---|
| Count | 598 |
| Mean | 0 days 07:07:49.966555183 |
| Std | 0 days 01:45:29.474139290 |
| Min | 0 days 00:06:00 |
| 25% | 0 days 06:14:30 |
| 50% | 0 days 07:24:00 |
| 75% | 0 days 08:10:45 |
| Max | 0 days 13:48:00 |

#### Average Coefficient of Variation (CV)

The CV was computed using the following formula:

$$CV = \frac{\text{Standard Deviation}}{\text{Mean}}$$

For some window sizes, some of the feature values were uniformly zero. This led to undefined CV, which are represented by grey boxes.

#### Heatmap of Average CV in active period

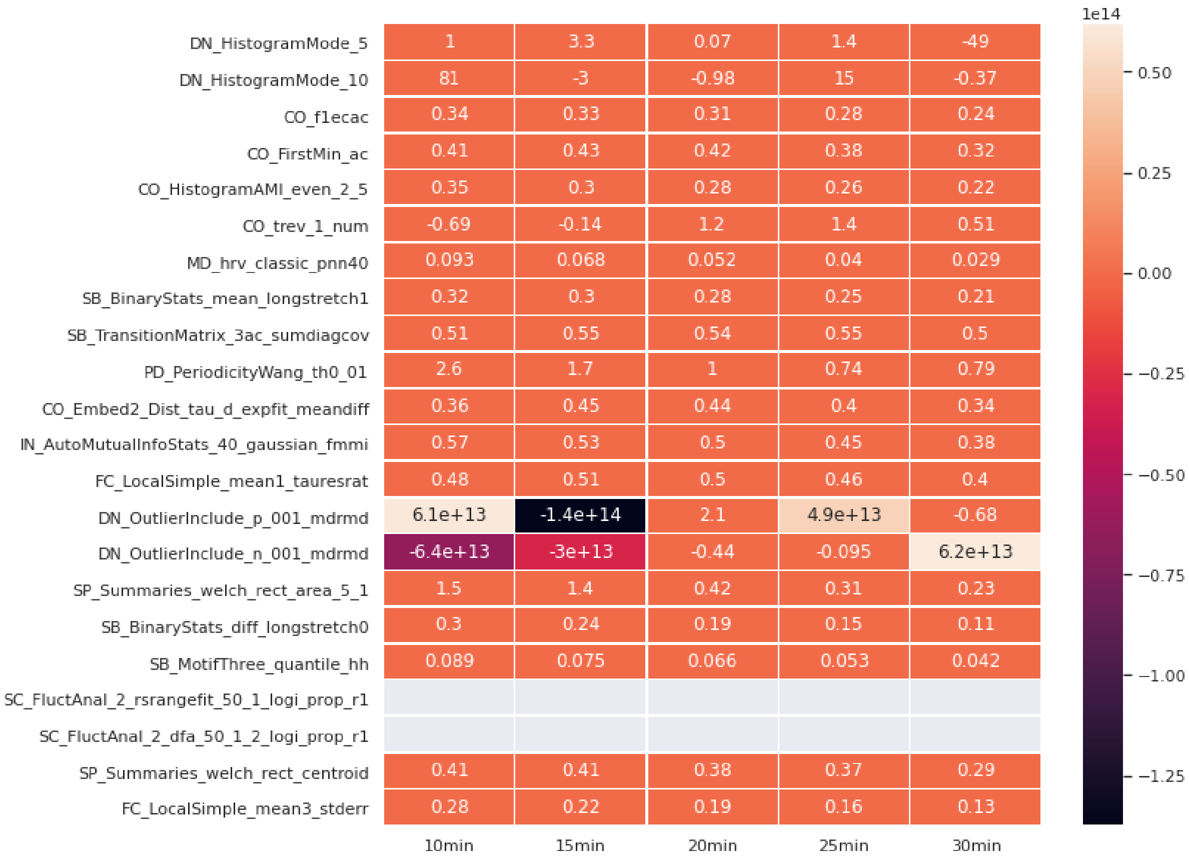

#### Heatmap of Average CV in sedentary period

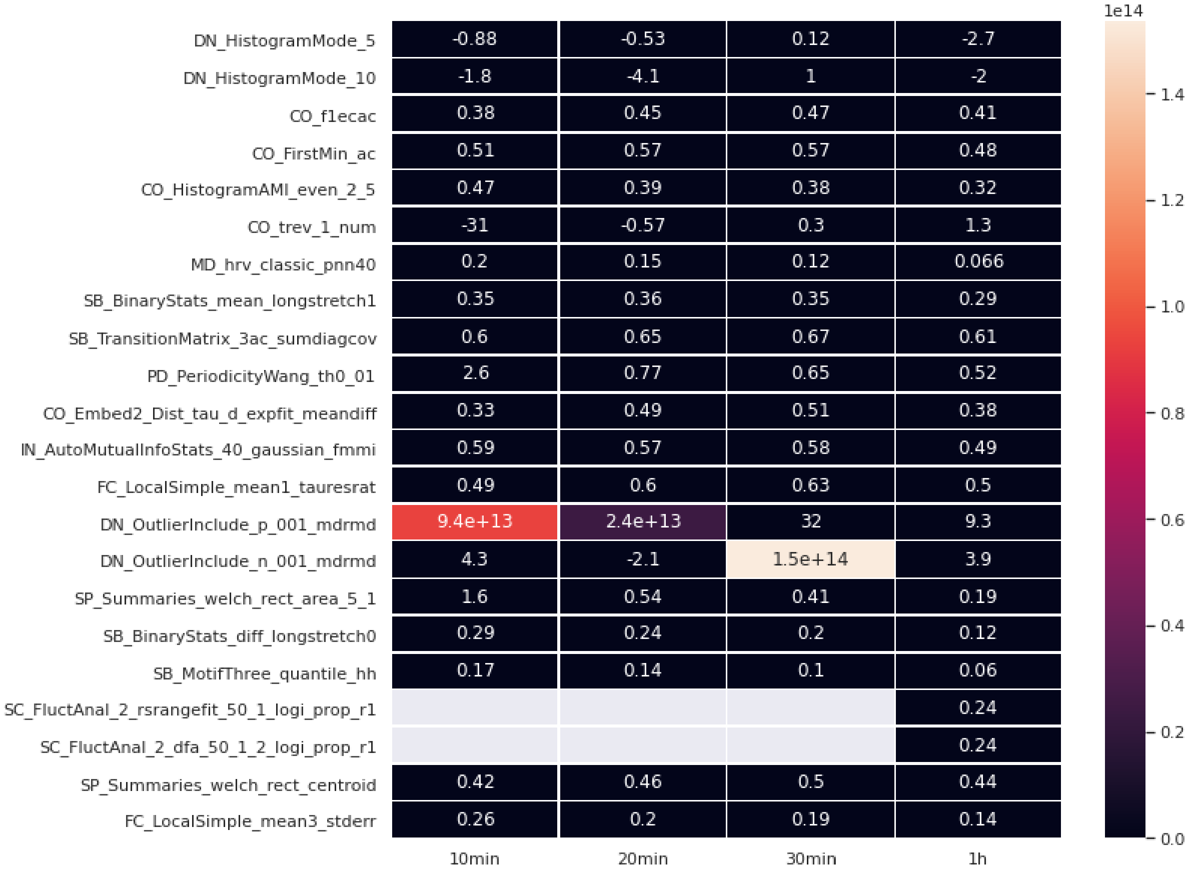

#### Heatmap of Average CV in sleep period

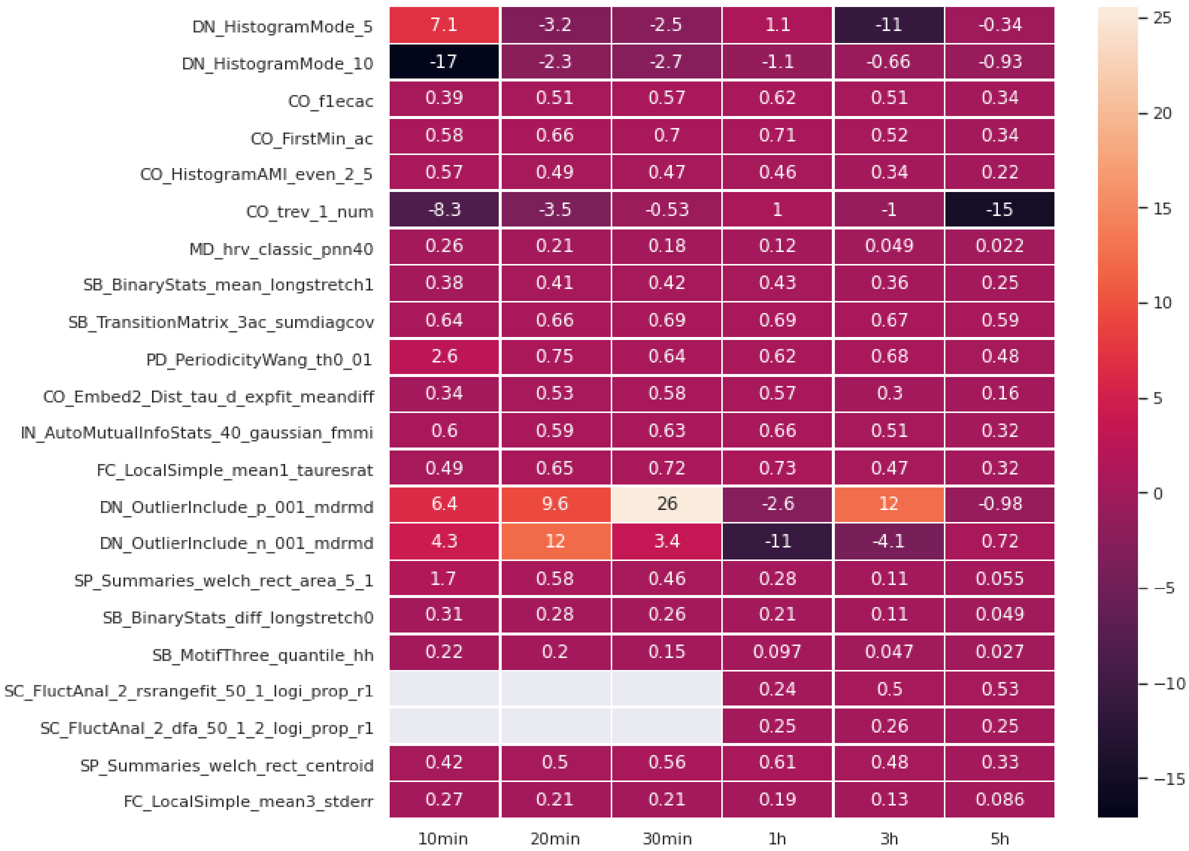

#### SI-2: Distribution of Cardiometabolic Risk Targets

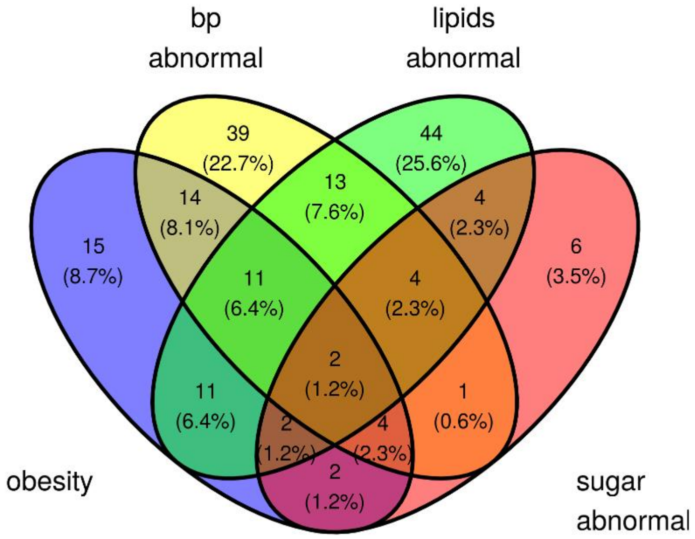

The training set for cardiometabolic risk targets consisted of 321 subjects. Of these, 172 subjects belong to at least one of the four major classes (obesity, blood pressure abnormalities, lipids abnormalities and sugar abnormalities). The distribution of these 172 subjects in the four classes are shown in the Venn diagram above.

Due to the extremely small number of subjects in the "sugar abnormal" class, we did not train any models for this class. However, the subjects of the "sugar abnormal" class are included in the higher-order class "anyRISKoutof9".

#### SI-3: PGS mapped trait ontology

##### Lipids Abnormality

| | PGS ID | Mapped Trait Ontology | Num. of Variants |
|---|---|---|---|
| 1 | PGS000060 | high density lipoprotein cholesterol measurement | 46 |
| 2 | PGS000061 | low density lipoprotein cholesterol measurement | 37 |
| 3 | PGS000062 | total cholesterol measurement | 52 |
| 4 | PGS000063 | triglyceride measurement | 32 |
| 5 | PGS000065 | low density lipoprotein cholesterol measurement | 103 |
| 6 | PGS000115 | low density lipoprotein cholesterol measurement | 223 |
| 7 | PGS000192 | high density lipoprotein cholesterol measurement | 9 |
| 8 | PGS000309 | high density lipoprotein cholesterol measurement | 247 |
| 9 | PGS000310 | low density lipoprotein cholesterol measurement | 194 |
| 10 | PGS000311 | total cholesterol measurement | 234 |
| 11 | PGS000340 | low density lipoprotein cholesterol measurement | 28 |
| 12 | PGS000677 | total cholesterol measurement | 17,204 |
| 13 | PGS000688 | low density lipoprotein cholesterol measurement | 16,184 |
| 14 | PGS000699 | triglyceride measurement | 16,003 |

##### Blood Pressure Abnormality

| | PGS ID | Mapped Trait Ontology | Num. of Variants |
|---|---|---|---|
| 1 | PGS000301 | systolic blood pressure | 970 |
| 2 | PGS000302 | diastolic blood pressure | 962 |

##### Obesity

| | PGS ID | Mapped Trait Ontology | Num. of Variants |
|---|---|---|---|
| 1 | PGS000298 | Body mass index | 941 |

### SI-4: Model Details for Genomic and Lifestyle Targets

#### Genomic Risk Targets

Threshold = 80/20$^{th}$ percentile

| Genomic Risk Targets | Number of Subjects with High Genomic Risk | Number of Subjects with Normal Genomic Risk |
|---|---|---|
| Lipids Abnormalities | 238 | 83 |
| Blood Pressure Abnormalities | 79 | 242 |
| Obesity | 69 | 252 |

Threshold = 85/15$^{th}$ percentile

| Genomic Risk Targets | Number of Subjects with High Genomic Risk | Number of Subjects with Normal Genomic Risk |
|---|---|---|
| Lipids Abnormalities | 220 | 101 |
| Blood Pressure Abnormalities | 67 | 254 |
| Obesity | 45 | 276 |

Threshold = 90/10$^{th}$ percentile

| Genomic Risk Targets | Number of Subjects with High Genomic Risk | Number of Subjects with Normal Genomic Risk |
|---|---|---|
| Lipids Abnormalities | 169 | 152 |
| Blood Pressure Abnormalities | 40 | 281 |
| Obesity | 33 | 288 |

#### Lifestyle Targets

| Target | Total Number of Subjects (Training Set) | Number of Class Labels |
|---|---|---|
| Pain.Discomfort | 398 | True: 71<br>False: 327 |
| Anxiety.Depression | 408 | True: 47<br>False: 361 |
| Stress.Level | 411 | Low: 149<br>Moderate: 180<br>High: 82 |
| Health.State | 411 | Low: 30<br>Moderate: 123<br>High: 258 |
| Alcohol.In3Mths | 407 | True: 148<br>False: 259 |
| High.Caffeine | 382 | True: 321<br>False: 61 |
| Relaxation.Therapies | 411 | True: 27<br>False: 384 |

### SI-5: Distribution of Catch22 Scores

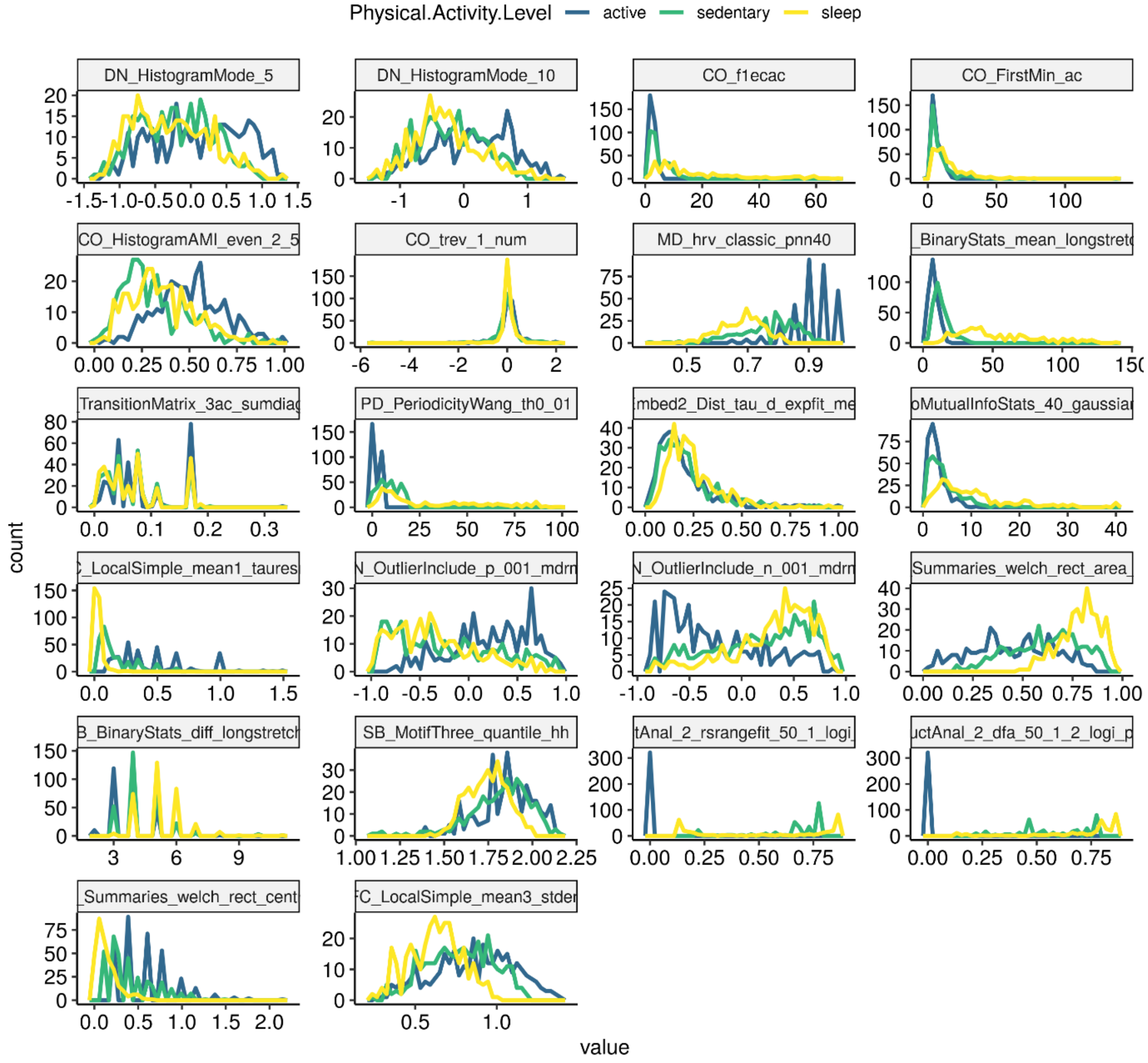

*Frequency polygons of the 22 high resolution features, coloured by the physical activity period they were derived from. These are all based on the training set data (*N* = 321).

#### SI-6: Sensitivity Analysis - Model Performance on Genomic Risk Targets

The PGS risk groups in Figure 6 of the main paper were defined by using the 90th (or 10th) percentile of the associated PGS as cut-offs. In order to determine if the obtained results were simply due to the defined cut-offs, we consider two other cut-offs and present the results below.

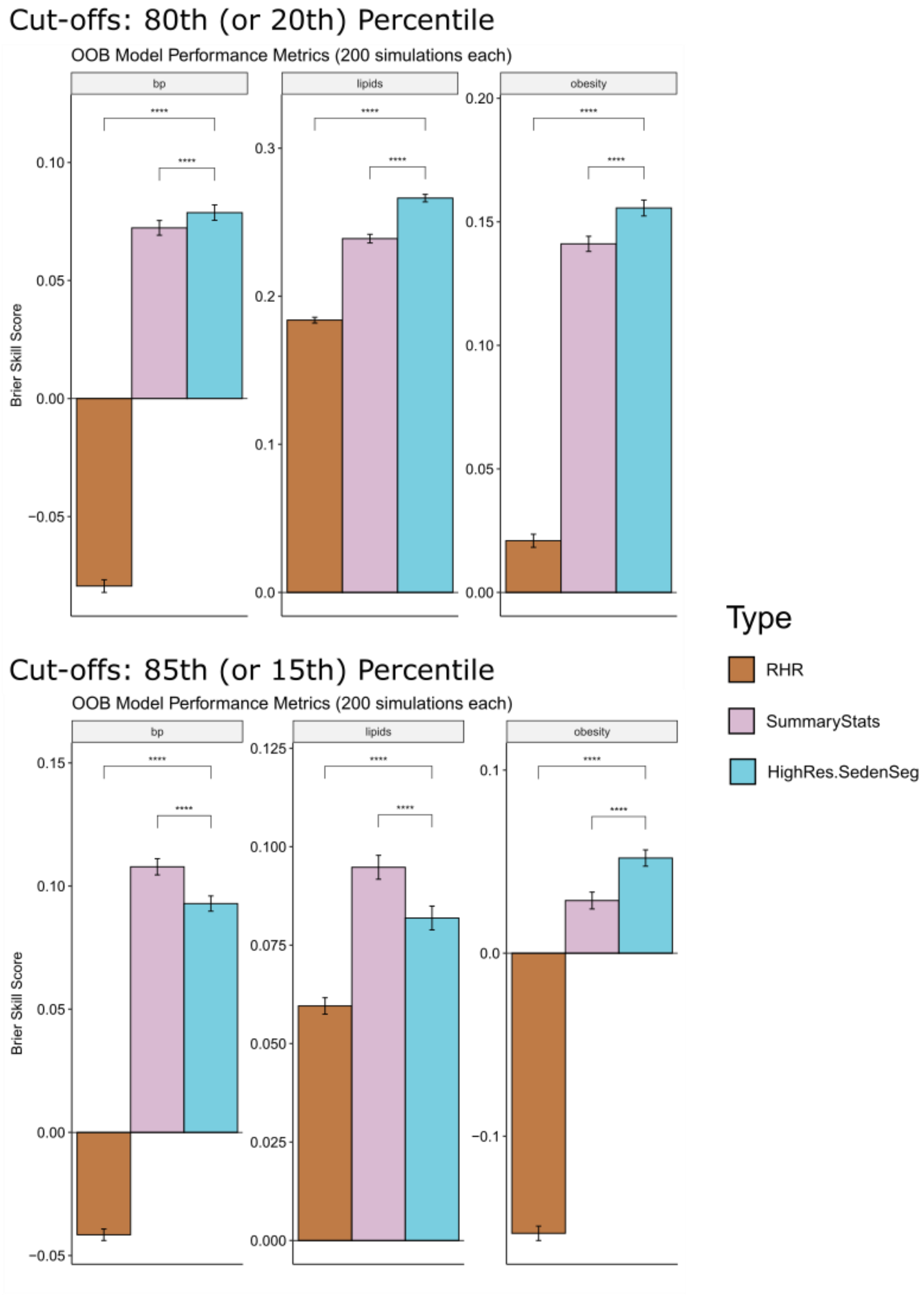

#### SI-7: Average Variable Importance of HighRes.SedenSeg Models

Variable importance are obtained as the average across 200 simulations. MDA: Mean Decrease in Accuracy.

**Target: anyRISKoutof9**

| | variable | MDA |
|---|---|---|
| 1 | Gender | 0.0145 |
| 2 | DN_HistogramMode_5.sedentary | 0.00522 |
| 3 | Age | 0.00472 |
| 4 | CO_Embed2_Dist_tau_d_expfit_meandiff.sedentary | 0.00137 |
| 5 | SB_BinaryStats_diff_longstretch0.sedentary | 0.00118 |
| 6 | CO_trev_1_num.sedentary | 0.00100 |
| 7 | DN_HistogramMode_10.sedentary | 0.000954 |
| 8 | DN_OutlierInclude_p_001_mdrmd.sedentary | 0.000813 |
| 9 | CO_HistogramAMI_even_2_5.sedentary | 0.000656 |
| 10 | SB_TransitionMatrix_3ac_sumdiagcov.sedentary | 0.000634 |

**Target: bp_abnormal**

| | variable | MDA |
|---|---|---|
| 1 | Age | 0.00895 |
| 2 | Gender | 0.00431 |
| 3 | CO_Embed2_Dist_tau_d_expfit_meandiff.sedentary | 0.00322 |
| 4 | CO_HistogramAMI_even_2_5.sedentary | 0.00194 |
| 5 | SC_FluctAnal_2_dfa_50_1_2_logi_prop_r1.sedentary | 0.00152 |
| 6 | FC_LocalSimple_mean1_tauresrat.sedentary | 0.00129 |
| 7 | PD_PeriodicityWang_th0_01.sedentary | 0.00128 |
| 8 | SP_Summaries_welch_rect_centroid.sedentary | 0.00108 |
| 9 | SB_TransitionMatrix_3ac_sumdiagcov.sedentary | 0.000810 |
| 10 | DN_HistogramMode_10.sedentary | 0.000284 |

**Target: lipids_abnormal**

| | variable | MDA |
|---|---|---|
| 1 | DN_HistogramMode_5.sedentary | 0.00504 |
| 2 | CO_trev_1_num.sedentary | 0.00407 |
| 3 | DN_OutlierInclude_n_001_mdrmd.sedentary | 0.00329 |
| 4 | DN_HistogramMode_10.sedentary | 0.00243 |
| 5 | CO_Embed2_Dist_tau_d_expfit_meandiff.sedentary | 0.00183 |
| 6 | PD_PeriodicityWang_th0_01.sedentary | 0.00181 |
| 7 | Gender | 0.00109 |
| 8 | DN_OutlierInclude_p_001_mdrmd.sedentary | 0.000989 |
| 9 | FC_LocalSimple_mean1_tauresrat.sedentary | 0.000962 |
| 10 | FC_LocalSimple_mean3_stderr.sedentary | 0.000925 |

**Target: obesity**

| | variable | MDA |
|---|---|---|
| 1 | DN_OutlierInclude_n_001_mdrmd.sedentary | 0.00505 |
| 2 | DN_HistogramMode_5.sedentary | 0.00190 |
| 3 | CO_Embed2_Dist_tau_d_expfit_meandiff.sedentary | 0.00171 |
| 4 | SB_MotifThree_quantile_hh.sedentary | 0.00127 |
| 5 | PD_PeriodicityWang_th0_01.sedentary | 0.00127 |

| | | |
|---|---|---|
| 6 | CO_FirstMin_ac.sedentary | 0.000552 |
| 7 | SC_FluctAnal_2_dfa_50_1_2_logi_prop_r1.sedentary | 0.000474 |
| 8 | SB_BinaryStats_mean_longstretch1.sedentary | 0.000294 |
| 9 | CO_trev_1_num.sedentary | 0.000292 |
| 10 | Gender | 0.0000236 |

#### SI-8: Additional Lifestyle Details for Illustrative CVD Subjects

##### Subject A

| Lifestyle Habits and Health Perceptions | Value |
|---|---|
| Pain.Discomfort | FALSE |
| Anxiety.Depression | FALSE |
| Stress.Level | Low |
| Health.State | Moderate |
| Alcohol.In3Mths | TRUE |
| High.Caffeine | TRUE |
| Relaxation.Therapies | FALSE |

| High Genetic Risk (based on $90^{th}$ percentile cut-off) | Value |
|---|---|
| Lipids | TRUE |
| Blood Pressure | TRUE |
| Obesity | TRUE |

##### Subject B

| Lifestyle Habits and Health Perceptions | Value |
|---|---|
| Pain.Discomfort | FALSE |
| Anxiety.Depression | FALSE |
| Stress.Level | Moderate |
| Health.State | Moderate |
| Alcohol.In3Mths | TRUE |
| High.Caffeine | TRUE |
| Relaxation.Therapies | FALSE |

| High Genetic Risk (based on $90^{th}$ percentile cut-off) | Value |
|---|---|
| Lipids | TRUE |
| Blood Pressure | TRUE |
| Obesity | FALSE |
